# Spatial Incidence of Noma in Northern Nigeria, 1999-2004: A model-based study

**DOI:** 10.1101/2025.06.19.25329958

**Authors:** Ramat Oyebunmi Braimah, Abdurrazaq Olanrewaju Taiwo, Seidu Bello, Mujtaba Bala, Azeez Butali, Bruno Oludare Ile-Ogedengbe, Abubakar Abdullahi Bello, Olawunmi Adedoyin Fatusi, Adetayo Oluwole Aborisade, John Adeoye

**Affiliations:** Department of Oral & Maxillofacial Surgery, Faculty of Dental Sciences, Usmanu Danfodiyo University, Sokoto, Nigeria; Cleft and Facial Deformity Foundation, Abuja, Nigeria; Department of Oral Pathology, Radiology and Medicine, College of Dentistry, University of Iowa, Iowa City, IA 52241, USA; Department of Dental & Maxillofacial Surgery, Federal Medical Centre, Birnin Kebbi, Kebbi, Nigeria; Noma Children’s Hospital, Sokoto, Nigeria; Department of Oral and Maxillofacial Surgery and Oral Pathology, Obafemi Awolowo University Teaching Hospitals Complex, Ile-Ife, Osun State, Nigeria; Department of Oral Diagnostic Sciences, Faculty of Dentistry, Bayero University, Kano, Nigeria; Division of Applied Oral Sciences and Community Dental Care, Faculty of Dentistry, University of Hong Kong, Hong Kong SAR, China

## Abstract

**Background:** Noma is a neglected tropical disease that affects the mouth and face, with a case fatality rate of 80-90%. Active disease is common among two to six-year-olds in Sub-Saharan Africa. The geographical distribution and risk of the disease are ill-defined, and incidence estimates for small areas are unavailable. This study aims to estimate the spatial incidence of noma in 296 small areas across 12 Northern Nigerian states using hierarchical Bayesian models to identify areas with high incidence risks.

**Methods:** This study used data on patients residing in Northern Nigeria managed at a major noma specialist center in Sokoto between 1999 and 2024 to estimate and map the small-area incidence risk of the disease in the region. Incidence was calculated dynamically at the local government area (LGA) level using the WHO Oral Health Unit’s method, while smoothed standardized incidence ratios (SIR) were modeled using a hierarchical Bayesian Poisson approach that accounts for spatial and non-spatial effects in risk estimation. Relative risks of noma incidence were used to draw choropleth maps followed by spatial autocorrelation analysis to determine clustering or variations in noma incidence among LGAs.

**Results:** All states had at least one LGA with a significantly higher noma incidence risk than the regional average. Notably, 20 LGAs in Sokoto (87%), 12 LGAs in Zamfara (85.7%), and 13 LGAs in Kebbi (61.9%) had significantly higher median SIRs than the regional average. Based on the relative risk ranks, Illela (SIR: 17.33 [16.64-18.04]), Wamako (SIR: 13.92 [13.36-14.48]), Goronyo (SIR: 11.66 [11.15-12.17]), and Tangaza (SIR: 11.09 [10.47-11.75]), LGAs in Sokoto state and Bade LGA (SIR: 12.05 [11.47-12.64]) in Yobe state had higher incidence risks than other LGAs in the study period. Autocorrelation analysis also confirmed the clustering of high noma incidence in northwest Nigeria involving 24 LGAs in Kebbi, Sokoto, and Zamfara states. Differences in noma incidence risks were also observed according to different sexes, age groups, and time intervals in the LGAs.

**Conclusions:** Spatial modeling was successful in highlighting small areas with high noma incidence risks in Northern Nigeria, which included at least one LGA in 12 northern Nigerian states. Maps of overall/stratified incidence risk estimates and trends should be considered to guide targeted individual and community actions that promote noma prevention/early detection to mitigate disease burden in Northern Nigeria.

## INTRODUCTION

Noma (cancrum oris) is a severe, rapidly progressing, and gangrenous neglected tropical disease that involves both the soft and hard tissues of the mouth and face ^1^. The disease often affects children between two and six years in highly vulnerable populations, with a case fatality rate of about 80-90% if untreated ^2,3^. However, survivors of active disease in childhood are left with extensive facial disfigurement, disability, stigma, and poor quality of life. Noma often starts as an elusive non-destructive inflammation of the gums (stage 0) and progresses rapidly through acute necrotizing ulcerative gingivitis (ANUG, stage 1) characterized by painful bleeding gums and fetor oris, edema (stage 2), gangrene (stage 3) before scarring (stage 4) and sequelae (stage 5) ^4^. Stages 0 to 2 are marked by reversible effects to the orofacial soft and hard tissues that are treatable by addressing risk factors and with antibiotics. The etiology of noma is multifactorial, often involving an interplay between several risk factors such as extreme poverty, malnutrition, poor general and oral hygiene, living nearby livestock, infectious diseases like HIV infection, malaria, measles, and immune compromising conditions resulting in microbial imbalance, thriving of opportunistic pathogens, and noma onset ^5–7^.

While noma was historically reported worldwide, it is presently viewed as a disease associated with impoverished communities. Incidence and prevalence of noma are higher in Sub-Saharan Africa (especially West Africa) than in any other region worldwide, where noma is a disease of public health concern ^3,5,7^. Incidence estimates of the disease in Sub-Saharan Africa range from 8.3 cases per 100,000 in north-central Nigeria to 16 cases per 100,000 in Ethiopia ^8,9^. Moreover, recent calculations estimated a 26-year noma incidence of 87.8 cases per 100,000, an annual incidence of 0.2 to 16.6 cases per 100,000 and an increasing incidence from 2020 onwards in Northern Nigeria ^10^.

Noma is frequently reported within Nigeria in the Northern geopolitical zones of the country ^3,11^. The incidence and prevalence of noma does exhibit significant geographical variability with different estimates obtained in studies conducted across large areas or states ^7,8,10^. Defining the geographical distribution and spatial incidence of noma represents an urgent research and public health need ^12^. If known, this will assist researchers, stakeholders, and policy makers in highlighting communities with high disease occurrence for targeted individual and community actions, guide inquiry into the rationale for disease incidence, and help formulate hypotheses on disease risk factors across different populations and locations. Therefore, this study aimed to estimate the small-area incidence of noma in northern Nigerian communities using hierarchical Bayesian models to identify at-risk areas. Additionally, a noma incidence atlas was developed for targeted individual and community actions in Northern Nigeria.

## METHODS

### Patient and population data

This study utilized data from patients with noma treated at the Noma Children’s Hospital (NCH), Sokoto from September 1999 to October 2024, for small-area incidence estimation and disease occurrence mapping. Demographics and clinical presentation of the patient cohort have been reported previously ^10^. Eligible patients included those with acute noma, i.e., stages 1 to 3 within the study period, who resided within any Northern Nigerian state at the time of presentation and diagnosis. As such, some patients living in the Niger Republic and southern Nigerian states at diagnosis were excluded. States in Northern Nigeria considered for case selection were Adamawa, Bauchi, Benue, Borno, Federal Capital Territory (FCT), Gombe, Jigawa, Kaduna, Kano, Katsina, Kebbi, Kogi, Kwara, Nasarawa, Niger, Plateau, Sokoto, Taraba, Yobe, and Zamfara. No age restrictions were applied as inclusion criteria, and patients without records of their state or local government area (LGA) of residence were excluded from this study. Overall, 916 NCH noma cases were used for incidence estimation and spatial mapping. Beyond the state of residence at diagnosis, other patient information collected for this cohort included their age, sex, year of diagnosis, and WHO noma stage at diagnosis ^13^.

Total population counts to calculate incidence ratios in each LGA were from the 2006 Population and Housing Census of Nigeria, which represents the most recent and official reference population in the country ^14^. For analysis by age categories (i.e., <5 years, 5-9 years, 10-17 years, and 18-39 years) and sex, specific reference counts for that population were used for incidence calculation and the determination of observed and expected counts during spatial modeling.

### Incidence estimation

This study estimated the incidence of noma (observed count) for each LGA following the expert consultation report of the WHO Oral Health Unit conducted using the Delphi method ^15^. LGA was chosen as the smallest geographic unit for incidence analysis since most cases had this information rather than township or ward-level information. Also, shapefiles of Nigerian areas were available at this level. Since only small-area incidence was calculated for Northern Nigeria as per study aims, we excluded states without eligible cases at NCH within the study period (i.e., Benue, FCT, Gombe, Kogi, Kwara, Nasarawa, Plateau, and Taraba). Incidence calculation involved three stages that estimate noma incidence as a function of the proportion of surviving cases from each state represented by NCH’s data, patient’s distance from their state of residence to NCH, and case mortality of noma ^8,10^.

In detail, since noma is a rapidly progressing disease, we first calculated the total number of surviving cases (*S*) as the ratio of the number of cases in NCH’s data (*N*) per LGA and the estimated proportion of surviving cases that were referred to the center (*p*). This was given by the equation: 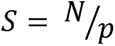 (where *p* is a value between 0 and 1).

Furthermore, *p* was dynamically selected according to the distance between the case’s state of residence and NCH following previous studies and expert opinions ^8–10^. For Sokoto state, approximately 20% of surviving cases are referred to NCH and for every 100km distance between other states and Sokoto, we reduced the proportion of surviving cases by 1%. Specific values of *p* (for each state) used for incidence estimation are available in **Supplementary Table 1**. Last, the incidence of noma was calculated for each LGA as the ratio of the total number of surviving cases afterward (*S*) and noma case survival rate of 10%.

### Bayesian spatial modeling of standardized incidence ratios

Estimated noma incidence calculated for 296 LGAs across 12 states was used for spatial analysis based on a Bayesian hierarchical (or fully) Poisson model (HBM) that assigns prior and hyperprior distributions to model parameters which account for within and between area variations in incidence ratios. Bayesian approach was favored over crude standardized incidence ratio (SIR) or relative risk calculations since these methods are often influenced by the size of the expected values and can produce an SIR of zero in geographic units without observed counts. Also, the Bayesian method allows for the estimation of smoothed incidence ratios, 95% credible intervals around point estimates, and posterior exceedance probability of point values being above the Northern Nigerian average population risk (i.e., SIR of 1). The HBM framework supports spatial dependence and often incorporates a conditional autoregressive (CAR) prior distribution as random effects that account for correlation between neighboring areas.

The theoretical foundations of our approach to spatial SIR calculation are presented in **Supplementary Methods**. In summary, we first calculated the crude SIR as the ratio of the count of observed noma cases (incidence estimates) to the expected number of cases for each LGA. The expected number of cases in each small area is the product of the population in each LGA and values obtained from the ratio of the sum of noma cases (observed count) to the sum of reference population in all 296 LGAs. Afterward, the incidence ratios were fitted using the Besag, York, and Mollie approach where random effects were specified using CAR priors in the HBM ^16^. The log SIR (*θ*) of an LGA (*i*) in the HBM model is specified as:

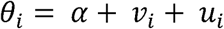

The overall risk effect (*α*) in the HBM was assigned an uninformative normal distribution with a mean *(μ)* of 0 and variance (*σ*^2^) of 10^6^. Further, non-spatial (unstructured) random effect (*v*) was modeled with a normal distribution, while spatial random effect (*u*) was modeled with an intrinsic CAR prior distribution. Within the CAR prior, a binary neighborhood matrix was specified to denote model proximity between small areas. Rook-adjacency spatial contiguity matrix was created using first-order weights to define areas that share a common boundary. Hyperpriors of spatial and unstructured random effects were vague gamma distributions selected following sensitivity analysis.

The HBM method was implemented using Gibbs sampling Markov chain Monte Carlo (MCMC) algorithm. Only a single Markov chain was run, and convergence of this chain was assessed using trace, density, and autocorrelation function plots and Monte Carlo (MC) error. For incidence risk estimates, 100,000 iterations were performed, and convergence of all nodes was achieved below 50,000 iterations. As such, the first 50,000 iterations served as burn-in, and we only sampled incidence ratios from the 50,000 iterations that followed at a thinning interval of 10 to ensure independence of estimates. Median (smoothed) SIRs of noma and their 95% credible intervals were obtained for 296 LGAs using an iteration sample size of 5,000. Spatial modeling was conducted using WINBUGS 1.4 *(MRC Biostatistics Unit, Cambridge, UK)*.

### Disease mapping

Median (smoothed) SIRs were used to draw choropleth maps indicating noma incidence risk in the 296 LGAs of 12 Northern Nigerian states between 1999 and 2024. Likewise, maps were created based on noma incidence ratios for different sexes, age groups, and the noma gangrene stage to showcase disease burden. All median SIR maps were accompanied by risk exceedance maps to showcase significant LGAs with high, same, or low incidence risk estimates as the regional average based on the distribution of the 95% credible interval. Moreover, graphs showcasing the different median SIRs per five years of the study period were also drawn for the LGAs at the state level. Nigerian LGA shapefiles and geometry data were obtained from the Humanitarian Data Exchange platform of the UN Office for the Coordination of Humanitarian Affairs (OCHA), Nigeria ^17^. Shapefiles were modified according to our data using QGIS ^18^ to include the 296 Northern Nigerian LGAs from 12 states in which incidence risks were calculated.

Spatial clustering and variation were assessed using Global Moran’s I and local indicator of spatial autocorrelation (LISA) tests. Only LGAs that had smoothed estimates and 95% credible intervals risk that indicated high or low noma incidence were used to define spatial autocorrelation. Also, probability values of spatial autocorrelation were determined using MC randomization at 99,999 permutations. Median SIR/significance mapping and spatial autocorrelation analyses were performed in GeoDa v 1.20 ^19^.

### Ethics

Ethical approval to conduct the study was granted by the Sokoto State Ministry of Health Ethics Research Committee with reference number SKHREC/037/2024. Informed consent was waived due to the retrospective nature of this study.

## RESULTS

### Incidence estimation for areal noma data

Demographic characteristics and clinical presentation of 916 noma cases at NCH are detailed in **Supplementary Table 2**. Overall, 51,048 new noma cases were estimated based on NCH’s data between 1999 and 2024. Also, the estimated 26-year incidence for the 296 LGAs across 12 Northern Nigerian states is in **Supplementary Table 3**. Overall, we calculated that 188 LGAs had little to no new noma cases (63.5%), while Gada LGA in Sokoto had the highest estimated number of new noma cases in the study period (n = 2600).

### Small-area noma incidence risk in Northern Nigeria

Sample trace and density plots showing convergence of the risk estimate nodes of the HBM are in **Supplementary Figures 1-2**. Also, autocorrelation function plots were close to zero for lags > 0, indicating independence and randomness of risk estimate sampling (**Supplementary Figure 3**). Choropleth map of the relative risk of noma incidence between 1999 and 2024 in Northern Nigeria and their significance levels are drawn in **Figure 1**. Overall, a majority of the LGAs (74.7%, 221) had median SIRs that were significantly below the average regional SIR of Northern Nigeria (i.e., SIR=1) while 21.6% (64) of LGAs had higher than the average SIR. Notably, at the state level, many LGAs in three north-western states i.e., 87% of LGAs in Sokoto (20), 85.7% of LGAs in Zamfara (12), and 61.9% of LGAs in Kebbi (13) had significantly higher relative risk of noma incidence than the average regional incidence risk (**Supplementary Figure 4**). While in north-east Nigeria, 17.6% of LGAs in Yobe state (3) had increased median SIRs than the regional average. At the LGA level, Illela (SIR: 17.33 [16.64-18.04]), Wamako (SIR: 13.92 [13.36-14.48]), Goronyo (SIR: 11.66 [11.15-12.17]), and Tangaza (SIR: 11.09 [10.47-11.75]), LGAs in Sokoto state and Bade LGA (SIR: 12.05 [11.47-12.64]) in Yobe state had the highest noma incidence risk within the study period. Detailed relative risk and 95% credible intervals indicating noma incidence for all 296 LGAs in Northern Nigeria are presented in **Supplementary Table 4**.

**Figure 1:**
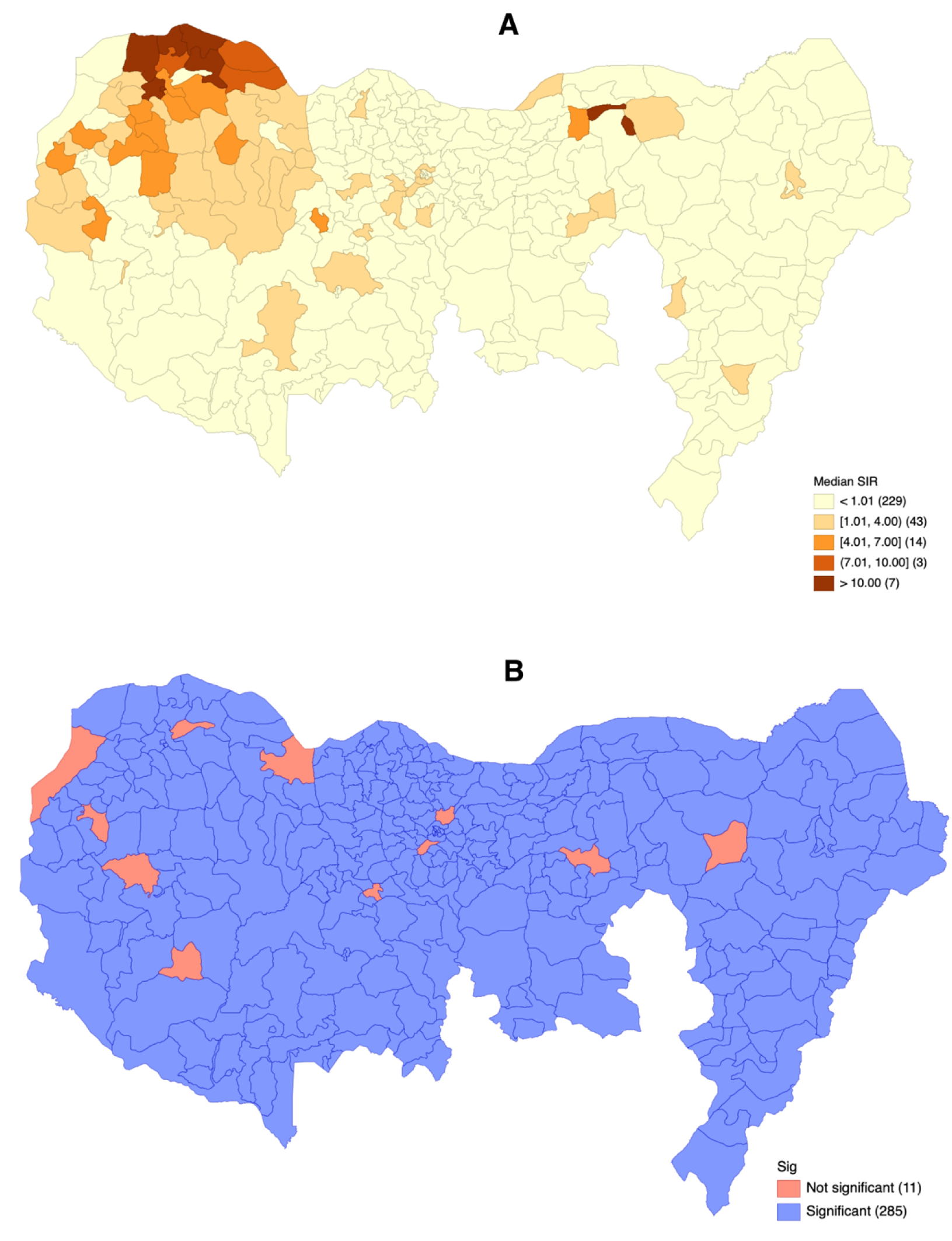
Estimated noma incidence risk in 296 Northern Nigerian local government areas (a) Map showing median SIR estimates distribution of noma for local government areas (b) Map showing the statistical significance of median SIR estimates based on the 95% credible intervals. (Areas with median SIRs and 95% credible intervals including 1 have the same risk level as the regional average while median SIRs below 1 have lower noma incidence risks compared to the regional average)

Upon removing estimated cases with ANUG and oedema from observed counts, the median SIR distribution for persons with gangrene (stage 3) was largely similar to the overall estimates (**Figure 2**). While Illela (SIR: 17.89 [17.17-18.62]), Wamako (SIR: 13.98 [13.40-14.58]), Bade (SIR: 13.54 [12.88-14.23]), and Goronyo (SIR: 11.47 [10.95-12.02]) still had very high incidence risks compared to the regional average as above, median SIR estimates for areas like Gada (SIR: 10.54 [10.10-10.98]) was higher for noma cases with gangrene when the LGAs were ranked based on risk levels. Though high, Tangaza LGA (SIR: 7.80 [7.26-8.38]) had a lower incidence risk for the gangrene stage of noma compared to the incidence risk for noma, irrespective of the stage.

**Figure 2:**
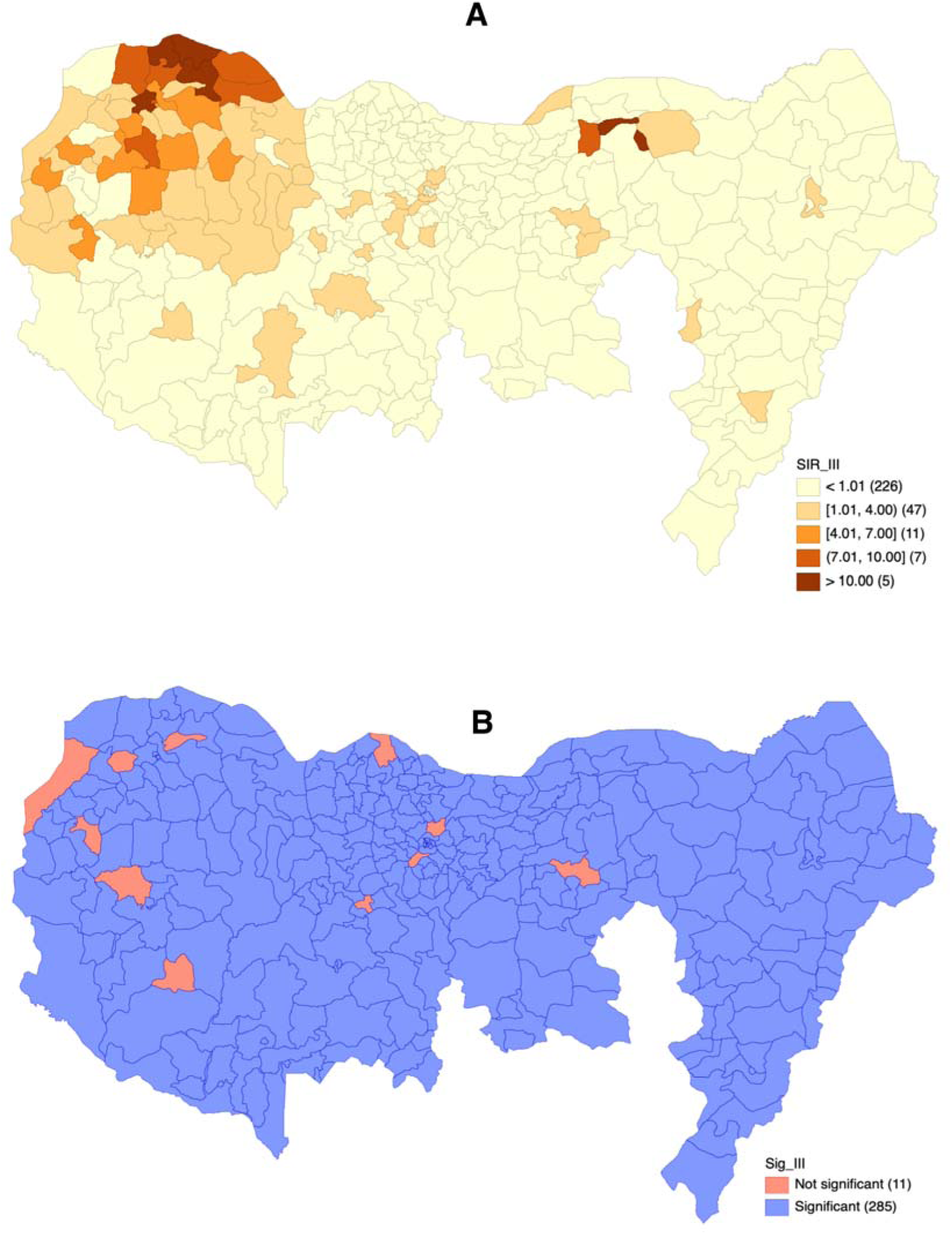
Estimated incidence risk of noma gangrene in 296 Northern Nigerian local government areas (a) Map showing median SIR estimates distribution of noma gangrene for local government areas (b) Map showing the statistical significance of median SIR estimates based on the 95% credible intervals. (Areas with median SIRs and 95% credible intervals including 1 have the same risk level as the regional average while median SIRs below 1 have lower noma incidence risks compared to the regional average)

Global Moran’s I (GMI) scatter plot for the median SIR estimates is in **Figure 3**. Overall, the relative risk of noma incidence in the LGAs displayed a positive spatial autocorrelation (GMI: 0.631, p<0.001), indicating that areas with similar noma incidence risks are often closer together and clustered. To further understand the relationship between areas and neighbors, LISA maps (**Figure 3**) showed evidence of clustering of 24 high-risk LGAs in Sokoto, Kebbi, and Zamfara of north-west Nigeria. Moreover, there were four significant LGAs with high noma incidence risk surrounded by low-risk areas (i.e., high-low spatial outliers), which included Bayo and Jere LGAs in Borno state, Rimi LGA in Katsina state, and Shiroro LGA in Niger state (**Figure 3**). Although Kudan LGA had the same risk as the average regional SIR based on the uncertainty around the estimates (i.e., 95% credible interval, **Supplementary Table 4**). Likewise, there were five significant LGAs with low noma incidence risk surrounded by high-risk areas (low-high spatial outliers), including Arewa-Dandi in Kebbi state, Gudu and Wurno in Sokoto state, and Jakusko and Karasuwa in Yobe state (**Figure 2**). However, Arewa-Dandi and Wurno had the same risk as the average regional SIR based on the 95% credible interval (**Supplementary Table 4**).

**Figure 3:**
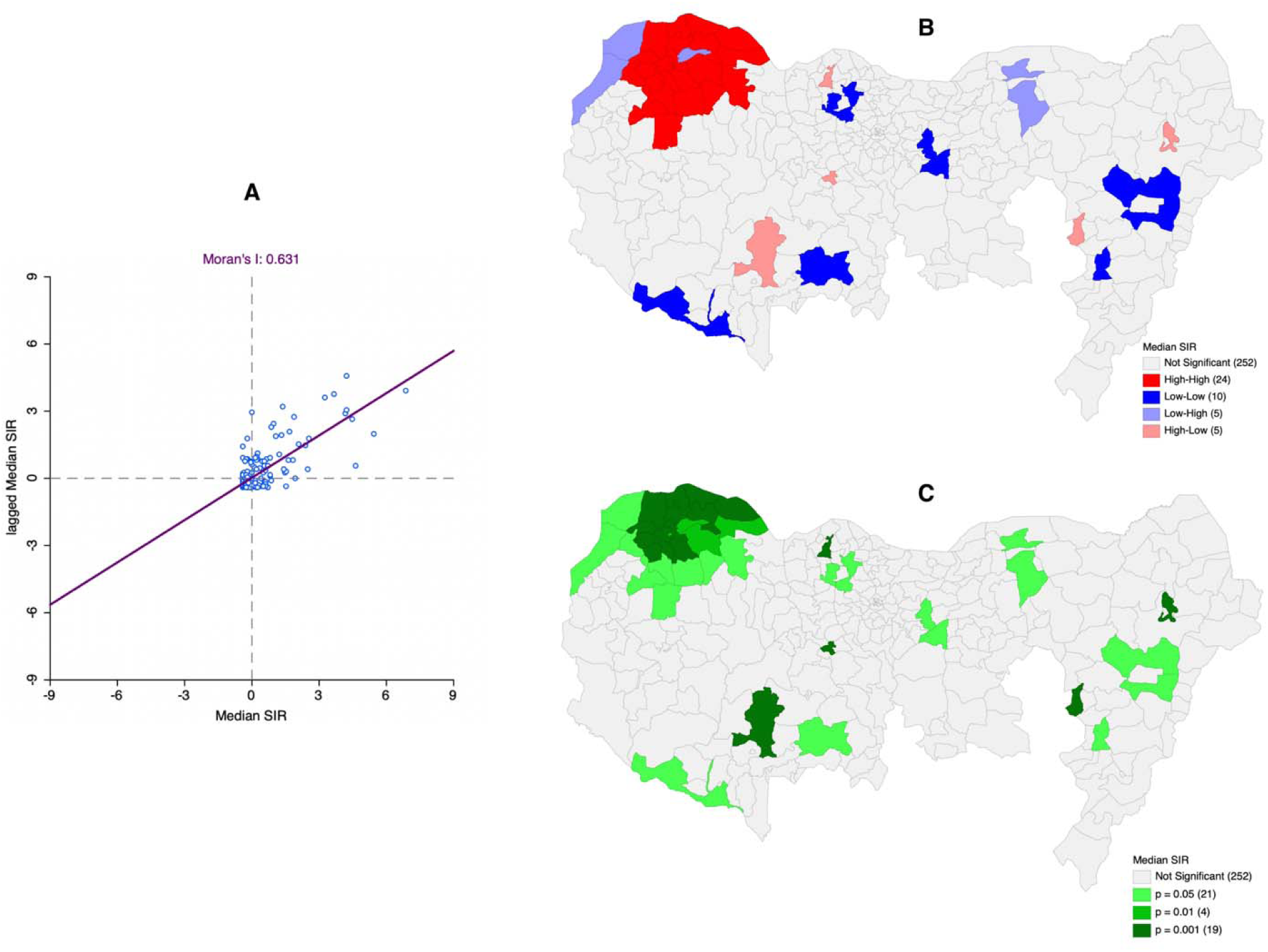
Spatial autocorrelation analysis (a) Scatter plot for global Moran’s I estimation of the median SIRs indicating positive autocorrelation in the noma incidence risks of the 296 LGAs (b) Local indicator of spatial association (LISA) maps showing spatial clusters and variations in noma incidence risk of 296 northern Nigerian LGAs (c) Significance maps for indicating noma incidence clusters.

### Stratified Noma Incidence Risk by Demographic Characteristics

Noma incidence risk maps by sex are in **Figure 4**, which indicated significant differences in the relative risk of noma incidence among males and females in certain LGAs. Notable areas with high noma incidence risk among females than males include Madobi and Minjibir in Kano, Bursari, Fakai, and Machina in Yobe, Guri (Jigawa), Igabi (Kaduna), Jere (Borno), Kalgo (Kebbi), Katagum (Bauchi), Mashi (Katsina), Shinkafi (Zamfara), Silame (Sokoto), and Yola North (Adamawa) (**Supplementary Table 5**). Likewise, LGAs with high noma incidence among males than females include Bauchi and Misau in Bauchi, Jega and Zuru in Kebbi, Dandume, Malumfashi, and Rimi in Katsina, Anka and Zurmi in Zamfara, Bayo (Borno), Damaturu (Yobe), Girei (Adamawa), Rano (Kano), Shiroro (Niger), and Yabo (Sokoto).

**Figure 4:**
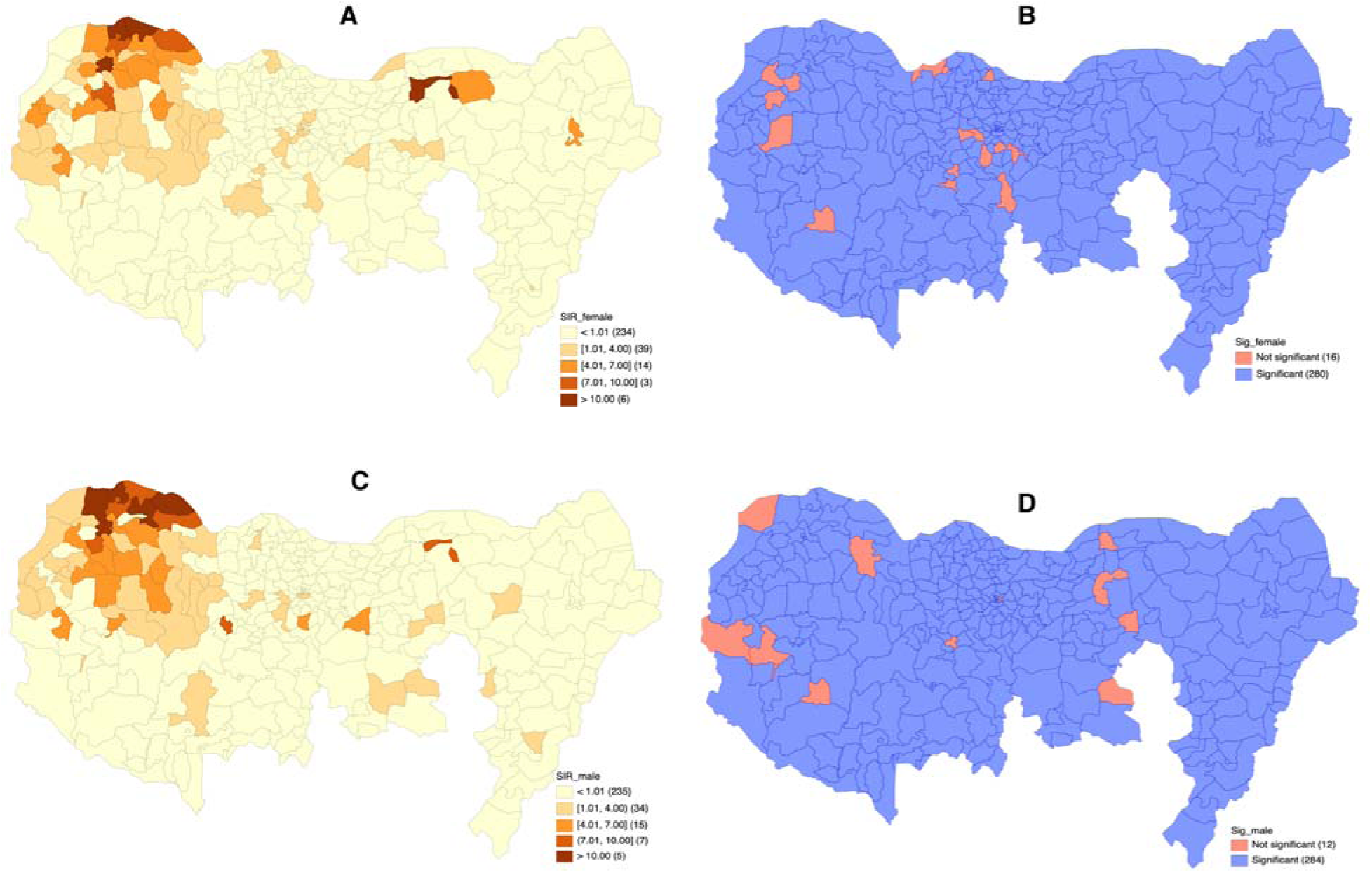
Stratified incidence risk of noma in 296 Northern Nigerian local government areas by sex (a) Map showing median SIR estimates distribution of noma among females (b) Map showing the statistical significance of median SIR estimates for females based on the 95% credible intervals (c) Map showing median SIR estimates distribution of noma among males (d) Map showing the statistical significance of median SIR estimates for males based on the 95% credible intervals.

Noma small-area incidence risk maps for different age groups in Northern Nigeria are in **Figure 5**. Overall, risk estimates higher than the regional average were seen in more LGAs for persons below ten years compared to adolescents and young adults. Tangaza (Sokoto) had the highest noma incidence risk among young adults, while Bade (Yobe) and Kware (Sokoto) had increased median SIR estimates for those aged 10 to 17 years. Bade LGA also had the highest incidence risk for children between 5 and 9 years, while Illela (Sokoto) had the highest incidence risk among children below five years old. Notably, LGAs like Gada, Sabon Birni, Gwadabawa, Sokoto North, Tambuwal, and Wamako in Sokoto, Bade in Yobe, and Maru in Zamfara had significantly higher incidence risk of noma among all four age categories in this study. Also, the spatial analysis revealed some LGAs like Bakura (Zamfara), Zaki (Bauchi), Kebbe and Wurno (Sokoto), Argungu (Kebbi), and Kano Municipal (Kano) that had high noma incidence risk estimates among young adults only.

**Figure 5:**
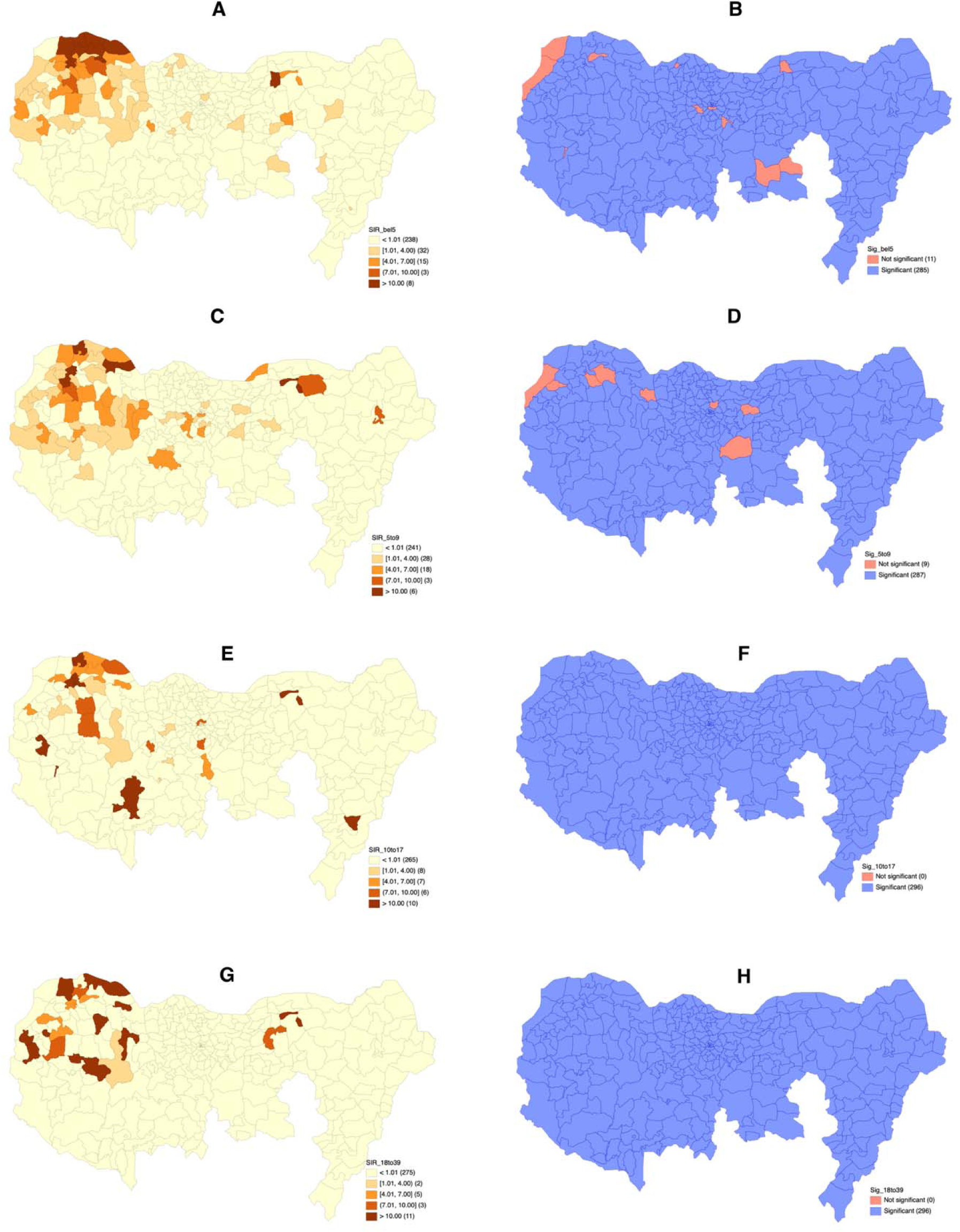
Stratified incidence risk of noma in 296 Northern Nigerian local government areas by age groups (a) Map showing median SIR estimates distribution of noma among children below five years old (b) Map showing the statistical significance of median SIR estimates for children below five years old based on the 95% credible intervals (c) Map showing median SIR estimates distribution of noma among 5-9 year olds (d) Map showing the statistical significance of median SIR estimates for 5-9 year olds based on the 95% credible intervals (e) Map showing median SIR estimates distribution of noma among 10-17 year olds (f) Map showing the statistical significance of median SIR estimates for 10-17 year olds based on the 95% credible intervals. (g) Map showing median SIR estimates distribution of noma among 18-39 year olds (h) Map showing the statistical significance of median SIR estimates for 18-39 year olds based on the 95% credible intervals.

### Temporal differences in noma spatial incidence

Differences in the noma incidence risk estimates at different time intervals for the 296 LGAs sampled are in **Supplementary Table 5 and Supplementary Figure 5**. Notably, only seven LGAs in Sokoto – Dange Shuni, Gada, Goronyo, Sabon Birni, Sokoto North, Tangaza, and Wamako, had consistently high noma incidence risk estimates at all time intervals of the study period. Considering the recent two time intervals (2015-2024), 60.9% of Sokoto LGAs (14), 47.6% of Kebbi LGAs (10), 21.4% of Zamfara LGAs (3), and 4.6% of Kano LGAs (Kiru and Kumbotso) had significantly increased relative risk of noma incidence than the regional average.

## DISCUSSION

This study calculated incidence risks of noma for 296 LGAs in twelve Northern Nigerian states using a fully Bayesian method and developed disease incidence maps that highlight high-risk areas in the region. Our analysis revealed that 64 LGAs had higher risks of noma incidence than the regional average, with at least one high-risk area in each of the twelve states sampled. Of note, many high-risk LGAs were in northwest Nigeria in the Sokoto-Zamfara-Kebbi axis, where a high incidence LGA cluster comprising 24 areas was defined. The top five high-risk LGAs based on overall incidence were Illela, Wamako, Goronyo, and Tangaza in Sokoto and Bade in Yobe, with four of these five LGAs also having high risks of stage 3 noma (gangrene). Stratified relative risks of noma incidence were also observed among the LGAs based on sex and age groups. Moreover, seven Sokoto LGAs (Dange Shuni, Gada, Goronyo, Sabo Birni, Sokoto North, Tangaza, and Wamako) had high incidence risk across all (five-year) time intervals in this study. In comparison, 29 more LGAs in Sokoto, Kebbi, Zamfara, and Kano had high noma incidence risks in the recent time intervals (2015-2024).

Before now, noma incidence calculations were available at the country, regional, or state level in Sub-Saharan Africa and Nigeria ^3,8–11,20^. This is the first study to estimate and define the geographical distribution of noma at the small-area level and identify spatial clusters of noma incidence risk in Northern Nigeria. The strengths of this study include employing a dynamic approach with the WHO Oral Health Unit’s method to estimate noma incidence and observed counts, implementing a comprehensive Bayesian framework for smoothing risk estimates according to spatial dependence, obtaining the uncertainty around the point risk estimates, and visualizing noma incidence risks using disease cartography methods that allow for spatial autocorrelation and significance analyses.

At least one LGA in the twelve Nigerian states studied had a high risk of noma incidence compared to the regional average. While the exact reasons may be unknown and represent areas for future scientific investigations, we offer some hypotheses and explanations. In north-east Nigeria, we observed that high-risk LGAs include areas that are susceptible to natural disasters like flooding and droughts. Flood-prone areas with low elevation and proximity to water bodies (like Benue, Yobe, Hadejia, and Misau rivers) highlighted from our analysis include Girei in Adamawa, Misau in Bauchi, Guri in Jigawa, and Bade in Yobe ^21^. Floods may result in detrimental effects on the soil, crops, and livestock, leading to food shortage, malnutrition, and noma susceptibility ^22^. Loss of occupation for farmers and cattle herders in these areas may also contribute to higher noma incidence. Water pollution and disease outbreaks due to flooding represent additional risk factors for noma ^23^. Moreover, areas with high incidence risks, like Damban and Misau in Bauchi and Bursari, Bade, and Machina in Yobe, have very hot climates with high ultraviolet indices and experience prolonged droughts that may lead to food shortages, compromised water quality and quantity, financial hardship, and challenged livelihoods, contributing to noma predisposition ^24–26^.

Though noma incidence in Nigeria is known to be highest in the northwestern region ^10,20^, our findings showed that disease distribution is higher among areas of Sokoto and Zamfara than in Kebbi state ^27^. This finding may be attributed to the poverty rate rankings in these states within the study period. However, examining high-risk LGA clusters in Sokoto, Kebbi, and Zamfara (like Illela, Goronyo, Tangaza, Sabon Birni, Isa, and Maradun) highlights that these areas include those most affected by banditry and insecurity in northwest Nigeria over the last two decades ^28^. This may have affected the agricultural occupations of residents or led to displacement from homes, culminating in vulnerable status and increased noma predisposition. This is further evident in the recent increase of noma incidence risk in areas like Shiroro of Niger state, where bandit attacks are frequent compared to other areas of the state ^29^.

Our results on the geographical distribution of noma incidence in Northern Nigeria does not support a hypothesis of increased incidence distribution among predominantly nomadic communities. Nonetheless, some high-risk LGAs (Girei in Adamawa, Jere in Borno, and Misau in Bauchi) with a large population of nomadic ethnicities such as the Fulani and Baggara Arabs were observed ^30^. Notably, we observed that the LGAs with higher noma incidence risks based on calculated overall, stratified, and time-interval estimates were closer to the Nigerian border with the Republic of Niger, especially in the regions of Tahoua, Maradi, and Dosso, which are among the poorest areas of the country with little agricultural yields, droughts, and food insecurity. Moreover, these regions have been highlighted as having high noma incidence and prevalence in Niger ^31^, which supports that national political borders alone do not define the geographic distribution of noma.

To employ the noma incidence maps developed in this study for designing and implementing prevention and early detection strategies in Northern Nigeria, we recommend that health workers, stakeholders, and policymakers use combined risk estimates (i.e., overall, stratified, and time interval) in formulating action plans and targeted programs for high-risk communities. Moreover, since areas with banditry, community displacement, and insecurity are likely to experience increased noma incidence, structuring future noma programs to target refugee and internally displaced persons camps will benefit noma prevention and early detection in Nigeria.

The study is not without limitations. First, we used a large hospital-based data to calculate the small-area incidence of noma in 296 local government areas of 12 Northern Nigerian states. While this study tried to adjust for the proportion of noma case presentation at the treatment center, it should be noted that our results are only estimates and should be interpreted as such when employing relative risks or noma incidence maps. Also, we have not modeled the incidence risk for eight states in Northern Nigeria due to little or ineligible data available for these areas at NCH. Last, spatial modeling could only provide information on high-risk areas without highlighting the rationale for risk levels. Given that these variables were not available in the NCH data for risk factor-based stratification, future studies should seek to explore predisposing factors in communities that may contribute to geographical distribution differences of noma incidence in Northern Nigeria.

## CONCLUSIONS

Bayesian spatial modeling was successful in estimating noma incidence risks for 296 LGAs in Northern Nigeria and highlighting areas with high disease incidence in the study period. These included at least one LGA in 12 northern Nigerian states. Clustering of high-risk LGAs was observed in the Sokoto-Zamfara-Kebbi parts of north-west Nigeria, while other high-risk LGAs (outliers) were observed in other northern Nigerian states. Areas with increased incidence risks mapped to LGAs with increased poverty, insecurity, hot climates, prolonged droughts, and flooding susceptibility. Disease incidence maps developed using estimated overall, stratified, and time interval-based incidence risk estimates should be considered when implementing targeted individual and community actions to promote noma prevention and early detection and mitigate disease burden in Northern Nigeria.

## Acknowledgements

The authors wish to thank the entire staff of the Noma Children’s Hospital, Sokoto for their efforts in providing comprehensive care for patients with noma in Nigeria.

## Data availability statement

Data for this study is presented in supplementary files of this article. Also, raw data may be obtained from the corresponding authors upon reasonable request. **Braimah RO** and **Bello AA** are guarantors for the NCH data used for incidence estimation and disease mapping.

## Code availability

Codes used to implement the fully Bayesian model in WINBUGS for sampling of posterior risk estimates are available: https://github.com/jaadeoye/Sokoto-HBM/blob/main/HBM.

## Author contributions

JA and ROB were involved in the conceptualization and design of this study. ROB, AOB, MB, BOI, and AAB contributed to data collection and curation. JA developed the methodology and performed all analyses in this study. SB and AB provided validation for the work. JA and ROB wrote the original draft of the manuscript. All authors critically reviewed the manuscript.

## REFERENCES

1. Baratti-Mayer D, Pittet B, Montandon D, Bolivar I, Bornand JE, Hugonnet S, Jaquinet A, Schrenzel J, Pittet D. Noma: an “infectious” disease of unknown aetiology. Lancet Infect Dis. 2003;3(7):419–31.

2. Feller L, Khammissa RAG, Altini M, Lemmer J. Noma (cancrum oris): An unresolved global challenge. Periodontol 2000. 2019;80(1):189–99.

3. Galli A, Brugger C, Fürst T, Monnier N, Winkler MS, Steinmann P. Prevalence, incidence, and reported global distribution of noma: a systematic literature review. Lancet Infect Dis. 2022;22(8):e221–e30.

4. Ashok N, Tarakji B, Darwish S, Rodrigues JC, Altamimi MA. A review on noma: a recent update. Global journal of health science. 2015;8(4):53.

5. Ogbureke KU, Ogbureke EI. NOMA: A Preventable “Scourge” of African Children. Open Dent J. 2010;4:201–6.

6. Adesola RO, Ajibade FA, Agaie MI. Noma (Cancrum oris) in Africa: A newly added neglected tropical disease. Rare. 2024;2:100031.

7. Verma A, Zaheer A, Ahsan A, Anand A, Abu Serhan H, Nazli Khatib M, Syed Zahiruddin Q, Gaidhane AM, Kukreti N, Rustagi S, Satapathy P, Sharma D, Arora M, Kumar Sharma R. Noma in the WHO’s list of neglected tropical diseases: A review of its impact on undeveloped and developing tropical regions. Prev Med Rep. 2024;43:102764.

8. Bello SA, Adeoye JA, Oketade I, Akadiri OA. Estimated incidence and Prevalence of noma in north central Nigeria, 2010-2018: A retrospective study. PLoS Negl Trop Dis. 2019;13(7):e0007574.

9. Gebretsadik HG. Estimation of the Prevalence of Noma in Ethiopia, 2007-2019: A Retrospective Study. Am J Trop Med Hyg. 2023;109(6):1388–92.

10. Braimah RO, Adeoye J, Taiwo AO, Bello S, Bala M, Butali A, Ile-Ogedengbe BO, Bello AA. Estimated incidence and clinical presentation of Noma in Northern Nigeria (1999-2024). PLoS Negl Trop Dis. 2025;19(5):e0012818.

11. Farley E, Oyemakinde MJ, Schuurmans J, Ariti C, Saleh F, Uzoigwe G, Bil K, Oluyide B, Fotso A, Amirtharajah M, Vyncke J, Brechard R, Adetunji AS, Ritmeijer K, van der Kam S, Baratti-Mayer D, Mehta U, Isah S, Ihekweazu C, Lenglet A. The prevalence of noma in northwest Nigeria. BMJ Glob Health. 2020;5(4):e002141.

12. Galli A, Comparet M, Dagne DA, Baratti-Mayer D, Cao TH, Guérin PJ, Guevara M, Hetzel MW, Jeantet C, Mpinga EK, Muendane V, Okanlawon M, Placella E, Ribes M, Sherlock M, Utzinger J, Steinmann P. Defining the noma research agenda. PLoS Negl Trop Dis. 2025;19(4):e0012940.

13. Khammissa RAG, Lemmer J, Feller L. Noma staging: a review. Trop Med Health. 2022;50(1):40.

14. Federal Republic of Nigeria. Report on the census 2006 final results. 2009 [cited 2024 December 12]. Available from: https://archive.gazettes.africa/archive/ng/2009/ng-government-gazette-dated-2009-02-02-no-2.pdf.

15. WHO Oral Health Unit. Noma today, a public health problem? 1998 [cited 2024 December 6]. Available from: https://iris.who.int/bitstream/handle/10665/63908/WHO_MMC_NOMA_98.1.pdf;jsessionid=A01101D5B4489A5859128E8DB5F81A64?sequence=1.

16. Besag J, York J, Mollié A. Bayesian image restoration, with two applications in spatial statistics. Annals of the institute of statistical mathematics. 1991;43:1–20.

17. The Humanitarian Data Exchange UN Office for the Coordination of Humanitarian Affairs (OCHA) Nigeria. Nigeria - Subnational Administrative Boundaries 2025 [cited 2025 June 9]. Available from: https://data.humdata.org/dataset/cod-ab-nga.

18. QGIS.org. QGIS Geographic Information System 2025 [cited 2025 June 9]. Available from: http://www.qgis.org/.

19. Anselin L, Syabri I, Kho Y. GeoDa: an introduction to spatial data analysis. Handbook of applied spatial analysis: Software tools, methods and applications: Springer; 2009. p. 73–89.

20. Fieger A, Marck KW, Busch R, Schmidt A. An estimation of the incidence of noma in north-west Nigeria. Trop Med Int Health. 2003;8(5):402–7.

21. Tudunwada IY, Abbas A. Geospatial Profiling of Flood Victims and Associated Losses of Lives and Properties in the Aftermath of 2022 Floods in Jigawa State, Northern Nigeria. Northern Nigeria.

22. Aldardasawi AM, Eren B. Floods and their impact on the environment. Academic Perspective Procedia. 2021;4(2):42–9.

23. Yang Z, Huang W, McKenzie JE, Yu P, Ju K, Wu Y, Wen B, Guo Y, Li S. Mortality and morbidity risks associated with floods: A systematic review and meta-analysis. Environmental Research. 2024;263:120263.

24. Dagona A, Dala A, Hassan A. Effects of Climate Change among Smallholder Farmers in Bade Local Government Area of Yobe State, Nigeria. Dutse Journal of Pure and Applied Sciences. 2024;10:55–66.

25. Eze JN. Drought occurrences and its implications on the households in Yobe state, Nigeria. Geoenvironmental Disasters. 2018;5(1):18.

26. Ja’afar Aliyu Umar KO, Hassan UM, Yusuf YY, Ahmed AH, Garba H. An Assessment of Groundwater Potential for Water Supply in Misau and Dambam Local Government Areas of Bauchi State, Nigeria. Applied Research and Innovation.2(2).

27. Pulse. Top 10 poorest states in Nigeria 2025 [cited 2025 June 18]. Available from: https://www.pulse.ng/articles/lifestyle/poorest-state-in-nigeria-2025050209332442160.

28. Punch Newspaper. Bandits’ control of 23 LGAs alarming 2023 [cited 2025 June 18]. Available from: https://punchng.com/bandits-control-of-23-lgas-alarming/.

29. Africa Independent Television. Facts About Shiroro, Niger Most Bandits Ridden Local Government Area 2024 [cited 2025 June 18]. Available from: https://ait.live/facts-about-shiroro-niger-most-bandits-ridden-local-government-area/.

30. Usman IS, Bakari U, Abdullahi A. Crop farmers and herders conflicts in Girei Local Government Area, Adamawa State, Nigeria: causes, repercussions and resolutions. Scientific Papers Series Management, Economic Engineering in Agriculture and Rural Development. 2017;17(1):467–72.

31. Issa AH, Ousmane KAK, Issa EOH, Shen J, Douma MD, Ibrahim AS, Eva M, Guan Y. Influencing factors for social acceptance of Noma (Cancrum Oris) patients in niger: A hospital-based cross-sectional study. Health. 2023;15(4):326–48.

