## Supplementary data for "Spatial Incidence of Noma in Northern Nigeria, 1999-2004: A model-based study"

**Supplementary Methods**

Besag, York, Mollie / Convolution CAR model

Taking the conventional standardized noma incidence ratio (*θ_h_*) which represents the risk of noma occurrence within non-overlapping geographic units (local government areas [LGAs] in 12 Northern Nigerian states) as the response variable ^1^:

^^^

n*_h_* [Σ y*_h_*]

[Σ n*_h_*]

*^h^*^=1^

^N^

^N^

*^h=^*^1^

*θ_h_*	=	*y_h_*	E*_h_* =

*E_h_*

{*h* = 1, 2, 3,…..,N}

Where *y_h_* represents the count of observed noma cases within respective LGAs ‘*h’* and *E_h_* is the expected number of noma cases within matched geographical areas which is calculated from the LGA population count at risk *n_h._* According to the hierarchical structure for disease mapping models described by Best et al ^2^:

^^^

*y_h_* ~ Poisson (e*^θh^*E*_h_)*

*_^_*

where *θ_h_* is the log relative risk in area *‘h’.*

*_^_*

*θ_h_ ~ α + η*

where *α* is the intercept/overall risk effect and *η* represents the random effect modelled by convolution priors. *α* was assigned vague normal distributions with mean *(μ)* 0 and variance *(σ^2^)* of 10^6^*_._*

*η =* υ*_h_* + ν*_h_*

where υ*_h_* and ν*_h_* represent the structured/spatial and unstructured/non-spatial random effects respectively. ν*_h_* is assigned a normal distribution while υ*_h_* is modelled using an intrinsic CAR prior for spatial effects.

ν*_h_* ~ *N*(0, τ_v_)

^2^

where τ^2^ is the inverse of the variance

1

Σ*_k_*ω*_hk_*

Σ

υ*_k_*ω*_hk_,*

*σ_u_*

^2^

Σ*_k_*ω*_hk_*

υ*_h_*|υ_-_*_h_* ~ *N*

Where ω*_hk_* represents a binary neighbourhood matrix to model the spatial closeness between the random effects.

value specified in the matrix if *h* and *k* DCDs are neighbours

1

ω*_hk_*

value specified in the matrix if *h* and *k* DCDs are not neighbours

0

Precision hyperparameters τ_v_ and τ_υ_ were also assigned uninformative gamma distributions. Different weakly informative priors were simulated to estimate these hyperparameters and sensitivity analyses was conducted to assess their individual effects on the smoothed risk estimates and the model’s goodness-of-fit. Hyperprior distributions finally selected following sensitivity analyses are given below:

*For Incidence model:*	τ_v_ ~ Γ(0.001, 0.001)

τ_υ_ ~ Γ(0.1, 0.1)

**References**

1.	Adeoye JA. New world vs old world: refining oral cancer screening, diagnosis, and prediction. HKU Theses Online (HKUTO). 2023.

2.	Best N, Richardson S, Thomson A. A comparison of Bayesian spatial models for disease mapping. Stat Methods Med Res 2005;14(1):35-9.

**Supplementary Table 1**

Proportion of surviving cases referred to Noma Children’s Hospital Sokoto for each state used to calculate the number of surviving cases

| **Distance rank** | **States** | **Average Distance to Sokoto (km)** | **Proportion of surviving cases,** $p$ |
| --- | --- | --- | --- |
| 0 | Sokoto | 0 | 0.2 |
| 1 | Kebbi | 150.9 | 0.1849 |
| 2 | Zamfara | 232.5 | 0.1768 |
| 3 | Katsina | 362 | 0.1638 |
| 4 | Niger | 497.6 | 0.1502 |
| 5 | Kano | 535 | 0.1465 |
| 6 | Kaduna | 542 | 0.1458 |
| 7 | Jigawa | 650.5 | 0.1349 |
| 8 | Yobe | 676 | 0.1324 |
| 9 | Bauchi | 823 | 0.1177 |
| 10 | Borno | 1080.5 | 0.92 |
| 11 | Adamawa | 1156.2 | 0.844 |

**Supplementary Table 2**

Demographic characteristics and clinical presentation of 916 noma patients used for incidence estimation and mapping

| **Variables** | | **N (%)** |
| --- | --- | --- |
| Age | Median (IQR) | 4 (3-7) |
| Age (category) | < 5 years | 518 (56.6) |
|  | 5 – 9 years | 253 (27.6) |
|  | 10 – 17 years | 89 (9.7) |
|  | 18 – 39 years | 51 (5.6) |
|  | 40 – 64 years | 5 (0.5) |
| Sex | Female | 422 (46.1) |
|  | Male | 494 (53.9) |
| State of residence at diagnosis | Adamawa | 3 (0.3) |
|  | Bauchi | 18 (2) |
|  | Borno | 6 (0.7) |
|  | Jigawa | 12 (1.3) |
|  | Kaduna | 21 (2.3) |
|  | Kano | 55 (6.0) |
|  | Katsina | 26 (2.8) |
|  | Kebbi | 112 (12.2) |
|  | Niger | 9 (1) |
|  | Sokoto | 506 (55.2) |
|  | Yobe | 27 (2.9) |
|  | Zamfara | 121 (13.2) |
| Year of diagnosis | 1999-2004 | 94 (10.3) |
|  | 2005-2009 | 54 (5.9) |
|  | 2010-2014 | 30 (3.3) |
|  | 2015-2019 | 120 (13.1) |
|  | 2020-2024 | 618 (67.5) |
| Stage of disease | ANUG | 5 (0.5) |
|  | Oedema | 101 (11.0) |
|  | Gangrene | 810 (88.4) |

**Supplementary Table 3**

Patient count, estimated incidence, and reference population by Local Government Areas

| **State** | **Local Government Area (LGA)** | **NCH patient count** | **Estimated Incidence / Observed Count** | **Reference Population**  **(2006 census)** |
| --- | --- | --- | --- | --- |
| Adamawa | Demsa | 0 | 0 | 178,407 |
| Adamawa | Fufore | 0 | 0 | 209,460 |
| Adamawa | Ganye | 0 | 0 | 169,948 |
| Adamawa | Girei | 2 | 237 | 129,855 |
| Adamawa | Gombi | 0 | 0 | 147,787 |
| Adamawa | Guyuk | 0 | 0 | 176,505 |
| Adamawa | Hong | 0 | 0 | 169,183 |
| Adamawa | Jada | 0 | 0 | 168,445 |
| Adamawa | Lamurde | 0 | 0 | 111,254 |
| Adamawa | Madagali | 0 | 0 | 135,142 |
| Adamawa | Maiha | 0 | 0 | 110,175 |
| Adamawa | Mayo-Belwa | 0 | 0 | 152,803 |
| Adamawa | Michika | 0 | 0 | 155,238 |
| Adamawa | Mubi North | 0 | 0 | 151,515 |
| Adamawa | Mubi South | 0 | 0 | 129,956 |
| Adamawa | Numan | 0 | 0 | 91,549 |
| Adamawa | Shelleng | 0 | 0 | 148,490 |
| Adamawa | Song | 0 | 0 | 195,188 |
| Adamawa | Toungo | 0 | 0 | 52,179 |
| Adamawa | Yola North | 1 | 118 | 199,674 |
| Adamawa | Yola South | 0 | 0 | 196,197 |
| Bauchi | Alkaleri | 0 | 0 | 328,284 |
| Bauchi | Bauchi | 4 | 340 | 493,730 |
| Bauchi | Bogoro | 0 | 0 | 83,809 |
| Bauchi | Damban | 4 | 340 | 150,212 |
| Bauchi | Darazo | 0 | 0 | 249,946 |
| Bauchi | Dass | 0 | 0 | 90,114 |
| Bauchi | Gamawa | 0 | 0 | 284,411 |
| Bauchi | Ganjuwa | 0 | 0 | 278,471 |
| Bauchi | Giade | 0 | 0 | 156,022 |
| Bauchi | Itas/Gadau | 0 | 0 | 228,527 |
| Bauchi | Jama'are | 0 | 0 | 117,482 |
| Bauchi | Katagum | 3 | 255 | 293,020 |
| Bauchi | Kirfi | 1 | 85 | 145,636 |
| Bauchi | Misau | 4 | 340 | 261,410 |
| Bauchi | Ningi | 1 | 85 | 385,997 |
| Bauchi | Shira | 0 | 0 | 233,999 |
| Bauchi | Tafawa-Balewa | 0 | 0 | 221,310 |
| Bauchi | Toro | 0 | 0 | 346,000 |
| Bauchi | Warji | 0 | 0 | 114,983 |
| Bauchi | Zaki | 1 | 85 | 189,703 |
| Borno | Abadam | 0 | 0 | 100,065 |
| Borno | Askira/Uba | 0 | 0 | 143,313 |
| Borno | Bama | 0 | 0 | 270,119 |
| Borno | Bayo | 1 | 109 | 79,078 |
| Borno | Biu | 0 | 0 | 175,760 |
| Borno | Chibok | 0 | 0 | 66,333 |
| Borno | Damboa | 0 | 0 | 233,200 |
| Borno | Dikwa | 0 | 0 | 105,042 |
| Borno | Gubio | 0 | 0 | 151,286 |
| Borno | Guzamala | 0 | 0 | 95,991 |
| Borno | Gwoza | 0 | 0 | 276,568 |
| Borno | Hawul | 0 | 0 | 120,733 |
| Borno | Jere | 5 | 543 | 209,107 |
| Borno | Kaga | 0 | 0 | 89,996 |
| Borno | Kala/Balge | 0 | 0 | 60,834 |
| Borno | Konduga | 0 | 0 | 157,322 |
| Borno | Kukawa | 0 | 0 | 203,343 |
| Borno | Kwaya Kusar | 0 | 0 | 56,704 |
| Borno | Mafa | 0 | 0 | 103,600 |
| Borno | Magumeri | 0 | 0 | 140,257 |
| Borno | Maiduguri | 0 | 0 | 540,016 |
| Borno | Marte | 0 | 0 | 129,409 |
| Borno | Mobbar | 0 | 0 | 116,633 |
| Borno | Monguno | 0 | 0 | 109,834 |
| Borno | Ngala | 0 | 0 | 236,498 |
| Borno | Nganzai | 0 | 0 | 99,074 |
| Borno | Shani | 0 | 0 | 100,989 |
| Jigawa | Auyo | 0 | 0 | 132,268 |
| Jigawa | Babura | 0 | 0 | 212,955 |
| Jigawa | Biriniwa | 0 | 0 | 142,015 |
| Jigawa | Birnin Kudu | 0 | 0 | 314,108 |
| Jigawa | Buji | 0 | 0 | 97,284 |
| Jigawa | Dutse | 0 | 0 | 251,135 |
| Jigawa | Gagarawa | 0 | 0 | 82,153 |
| Jigawa | Garki | 0 | 0 | 150,261 |
| Jigawa | Gumel | 0 | 0 | 106,371 |
| Jigawa | Guri | 10 | 741 | 113,363 |
| Jigawa | Gwaram | 0 | 0 | 271,368 |
| Jigawa | Gwiwa | 0 | 0 | 128,730 |
| Jigawa | Hadejia | 0 | 0 | 104,286 |
| Jigawa | Jahun | 1 | 74 | 229,882 |
| Jigawa | Kafin Hausa | 1 | 74 | 267,284 |
| Jigawa | Kaugama | 0 | 0 | 128,981 |
| Jigawa | Kazaure | 0 | 0 | 161,161 |
| Jigawa | Kiri Kasama | 0 | 0 | 192,583 |
| Jigawa | Kiyawa | 0 | 0 | 172,952 |
| Jigawa | Maigatari | 0 | 0 | 177,057 |
| Jigawa | Malam Madori | 0 | 0 | 164,791 |
| Jigawa | Miga | 0 | 0 | 127,876 |
| Jigawa | Ringim | 0 | 0 | 192,407 |
| Jigawa | Roni | 0 | 0 | 77,414 |
| Jigawa | Sule Tankarkar | 0 | 0 | 134,813 |
| Jigawa | Taura | 0 | 0 | 131,861 |
| Jigawa | Yankwashi | 0 | 0 | 95,643 |
| Kaduna | Birnin-Gwari | 0 | 0 | 258,581 |
| Kaduna | Chikun | 0 | 0 | 372,272 |
| Kaduna | Giwa | 0 | 0 | 292,384 |
| Kaduna | Igabi | 10 | 686 | 430,753 |
| Kaduna | Ikara | 0 | 0 | 194,723 |
| Kaduna | Jaba | 0 | 0 | 155,973 |
| Kaduna | Jema'a | 0 | 0 | 278,202 |
| Kaduna | Kachia | 0 | 0 | 252,568 |
| Kaduna | Kaduna North | 3 | 206 | 364,575 |
| Kaduna | Kaduna South | 2 | 137 | 402,731 |
| Kaduna | Kagarko | 0 | 0 | 239,058 |
| Kaduna | Kajuru | 0 | 0 | 109,810 |
| Kaduna | Kaura | 0 | 0 | 174,626 |
| Kaduna | Kauru | 0 | 0 | 221,276 |
| Kaduna | Kubau | 0 | 0 | 280,704 |
| Kaduna | Kudan | 2 | 137 | 138,956 |
| Kaduna | Lere | 0 | 0 | 339,740 |
| Kaduna | Makarfi | 0 | 0 | 146,574 |
| Kaduna | Sabon-Gari | 0 | 0 | 291,358 |
| Kaduna | Sanga | 0 | 0 | 151,485 |
| Kaduna | Soba | 0 | 0 | 291,173 |
| Kaduna | Zangon-Kataf | 0 | 0 | 318,991 |
| Kaduna | Zaria | 4 | 274 | 406,990 |
| Kano | Ajingi | 0 | 0 | 172,610 |
| Kano | Albasu | 0 | 0 | 187,639 |
| Kano | Bagwai | 0 | 0 | 161,533 |
| Kano | Bebeji | 1 | 68 | 191,916 |
| Kano | Bichi | 0 | 0 | 278,309 |
| Kano | Bunkure | 1 | 68 | 174,467 |
| Kano | Dala | 0 | 0 | 418,759 |
| Kano | Dambatta | 1 | 68 | 210,474 |
| Kano | Dawakin Kudu | 0 | 0 | 225,497 |
| Kano | Dawakin Tofa | 0 | 0 | 246,197 |
| Kano | Doguwa | 1 | 68 | 150,645 |
| Kano | Fagge | 0 | 0 | 200,095 |
| Kano | Gabasawa | 0 | 0 | 211,204 |
| Kano | Garko | 1 | 68 | 161,966 |
| Kano | Garum Mallam | 0 | 0 | 118,622 |
| Kano | Gaya | 0 | 0 | 207,419 |
| Kano | Gezawa | 0 | 0 | 282,328 |
| Kano | Gwale | 0 | 0 | 357,827 |
| Kano | Gwarzo | 1 | 68 | 183,624 |
| Kano | Kabo | 5 | 341 | 153,158 |
| Kano | Kano Municipal | 4 | 273 | 371,243 |
| Kano | Karaye | 0 | 0 | 144,045 |
| Kano | Kibiya | 0 | 0 | 138,618 |
| Kano | Kiru | 9 | 614 | 267,168 |
| Kano | Kumbotso | 10 | 683 | 294,391 |
| Kano | Kunchi | 0 | 0 | 110,170 |
| Kano | Kura | 0 | 0 | 143,094 |
| Kano | Madobi | 2 | 137 | 137,685 |
| Kano | Makoda | 0 | 0 | 220,094 |
| Kano | Minjibir | 3 | 205 | 219,611 |
| Kano | Nasarawa | 4 | 273 | 596,411 |
| Kano | Rano | 5 | 341 | 148,276 |
| Kano | Rimin Gado | 0 | 0 | 103,371 |
| Kano | Rogo | 0 | 0 | 227,607 |
| Kano | Shanono | 0 | 0 | 139,128 |
| Kano | Sumaila | 0 | 0 | 250,379 |
| Kano | Takai | 0 | 0 | 202,639 |
| Kano | Tarauni | 0 | 0 | 221,844 |
| Kano | Tofa | 0 | 0 | 98,603 |
| Kano | Tsanyawa | 0 | 0 | 157,730 |
| Kano | Tudun Wada | 0 | 0 | 228,658 |
| Kano | Ungogo | 7 | 478 | 365,737 |
| Kano | Warawa | 0 | 0 | 131,858 |
| Kano | Wudil | 0 | 0 | 188,639 |
| Katsina | Bakori | 0 | 0 | 149,516 |
| Katsina | Batagarawa | 0 | 0 | 189,059 |
| Katsina | Batsari | 0 | 0 | 207,874 |
| Katsina | Baure | 0 | 0 | 202,941 |
| Katsina | Bindawa | 0 | 0 | 151,002 |
| Katsina | Charanchi | 0 | 0 | 136,989 |
| Katsina | Dan Musa | 0 | 0 | 113,190 |
| Katsina | Dandume | 9 | 549 | 145,323 |
| Katsina | Danja | 0 | 0 | 125,481 |
| Katsina | Daura | 0 | 0 | 224,884 |
| Katsina | Dutsi | 0 | 0 | 120,902 |
| Katsina | Dutsin-Ma | 0 | 0 | 169,829 |
| Katsina | Faskari | 1 | 61 | 194,400 |
| Katsina | Funtua | 0 | 0 | 225,156 |
| Katsina | Ingawa | 0 | 0 | 169,148 |
| Katsina | Jibia | 1 | 61 | 167,435 |
| Katsina | Kafur | 0 | 0 | 209,360 |
| Katsina | Kaita | 0 | 0 | 182,405 |
| Katsina | Kankara | 0 | 0 | 243,259 |
| Katsina | Kankia | 0 | 0 | 151,395 |
| Katsina | Katsina | 3 | 183 | 318,132 |
| Katsina | Kurfi | 0 | 0 | 116,700 |
| Katsina | Kusada | 0 | 0 | 98,348 |
| Katsina | Mai'adua | 0 | 0 | 201,800 |
| Katsina | Malumfashi | 4 | 244 | 182,891 |
| Katsina | Mani | 0 | 0 | 176,301 |
| Katsina | Mashi | 2 | 122 | 171,070 |
| Katsina | Matazu | 0 | 0 | 113,814 |
| Katsina | Musawa | 0 | 0 | 170,006 |
| Katsina | Rimi | 4 | 244 | 154,092 |
| Katsina | Sabuwa | 0 | 0 | 140,679 |
| Katsina | Safana | 1 | 61 | 185,207 |
| Katsina | Sandamu | 1 | 61 | 136,944 |
| Katsina | Zango | 0 | 0 | 156,052 |
| Kebbi | Aleiro | 3 | 162 | 67,078 |
| Kebbi | Arewa-Dandi | 3 | 162 | 189,728 |
| Kebbi | Argungu | 2 | 108 | 200,248 |
| Kebbi | Augie | 6 | 324 | 116,368 |
| Kebbi | Bagudo | 5 | 270 | 238,014 |
| Kebbi | Birnin Kebbi | 21 | 1136 | 268,620 |
| Kebbi | Bunza | 10 | 541 | 123,547 |
| Kebbi | Dandi | 6 | 324 | 146,211 |
| Kebbi | Fakai | 2 | 108 | 119,772 |
| Kebbi | Gwandu | 4 | 216 | 151,077 |
| Kebbi | Jega | 3 | 162 | 197,757 |
| Kebbi | Kalgo | 4 | 216 | 84,928 |
| Kebbi | Koko/Besse | 15 | 811 | 154,818 |
| Kebbi | Maiyama | 1 | 54 | 173,759 |
| Kebbi | Ngaski | 0 | 0 | 126,102 |
| Kebbi | Sakaba | 0 | 0 | 91,728 |
| Kebbi | Shanga | 1 | 54 | 127,142 |
| Kebbi | Suru | 6 | 324 | 148,474 |
| Kebbi | Wasagu/Danko | 7 | 379 | 265,271 |
| Kebbi | Yauri | 4 | 216 | 100,564 |
| Kebbi | Zuru | 9 | 487 | 165,335 |
| Niger | Agaie | 0 | 0 | 132,098 |
| Niger | Agwara | 0 | 0 | 57,347 |
| Niger | Bida | 0 | 0 | 185,553 |
| Niger | Borgu | 0 | 0 | 172,835 |
| Niger | Bosso | 0 | 0 | 148,136 |
| Niger | Chanchaga | 0 | 0 | 202,151 |
| Niger | Edati | 0 | 0 | 159,818 |
| Niger | Gbako | 0 | 0 | 126,845 |
| Niger | Gurara | 0 | 0 | 90,879 |
| Niger | Katcha | 0 | 0 | 120,893 |
| Niger | Kontagora | 2 | 133 | 151,968 |
| Niger | Lapai | 0 | 0 | 117,021 |
| Niger | Lavun | 0 | 0 | 209,777 |
| Niger | Magama | 1 | 67 | 181,470 |
| Niger | Mariga | 0 | 0 | 199,600 |
| Niger | Mashegu | 0 | 0 | 215,197 |
| Niger | Mokwa | 0 | 0 | 242,858 |
| Niger | Muya | 0 | 0 | 103,461 |
| Niger | Paikoro | 0 | 0 | 158,178 |
| Niger | Rafi | 0 | 0 | 186,118 |
| Niger | Rijau | 1 | 67 | 176,199 |
| Niger | Shiroro | 5 | 333 | 235,665 |
| Niger | Suleja | 0 | 0 | 215,075 |
| Niger | Tafa | 0 | 0 | 83,874 |
| Niger | Wushishi | 0 | 0 | 81,756 |
| Sokoto | Binji | 7 | 350 | 104,274 |
| Sokoto | Bodinga | 11 | 550 | 174,302 |
| Sokoto | Dange Shuni | 15 | 750 | 193,443 |
| Sokoto | Gada | 52 | 2600 | 249,051 |
| Sokoto | Goronyo | 40 | 2000 | 182,118 |
| Sokoto | Gudu | 1 | 50 | 95,400 |
| Sokoto | Gwadabawa | 38 | 1900 | 231,569 |
| Sokoto | Illela | 49 | 2450 | 150,133 |
| Sokoto | Isa | 20 | 1000 | 150,268 |
| Sokoto | Kebbe | 1 | 50 | 123,154 |
| Sokoto | Kware | 14 | 700 | 134,084 |
| Sokoto | Rabah | 17 | 850 | 149,152 |
| Sokoto | Sabon Birni | 38 | 1900 | 207,470 |
| Sokoto | Shagari | 20 | 1000 | 156,907 |
| Sokoto | Silame | 6 | 300 | 104,601 |
| Sokoto | Sokoto North | 48 | 2400 | 233,012 |
| Sokoto | Sokoto South | 16 | 800 | 197,686 |
| Sokoto | Tambuwal | 23 | 1150 | 225,917 |
| Sokoto | Tangaza | 24 | 1200 | 114,770 |
| Sokoto | Tureta | 5 | 250 | 68,414 |
| Sokoto | Wamako | 47 | 2350 | 179,246 |
| Sokoto | Wurno | 3 | 150 | 162,403 |
| Sokoto | Yabo | 11 | 550 | 115,302 |
| Yobe | Bade | 21 | 1586 | 139,804 |
| Yobe | Bursari | 3 | 227 | 109,692 |
| Yobe | Damaturu | 1 | 76 | 87,706 |
| Yobe | Fika | 0 | 0 | 136,736 |
| Yobe | Fune | 0 | 0 | 301,954 |
| Yobe | Geidam | 0 | 0 | 155,740 |
| Yobe | Gujba | 0 | 0 | 129,797 |
| Yobe | Gulani | 0 | 0 | 103,516 |
| Yobe | Jakusko | 0 | 0 | 232,458 |
| Yobe | Karasuwa | 0 | 0 | 105,514 |
| Yobe | Machina | 1 | 76 | 60,994 |
| Yobe | Nangere | 0 | 0 | 87,517 |
| Yobe | Nguru | 1 | 76 | 150,699 |
| Yobe | Potiskum | 0 | 0 | 204,866 |
| Yobe | Tarmua | 0 | 0 | 77,667 |
| Yobe | Yunusari | 0 | 0 | 125,940 |
| Yobe | Yusufari | 0 | 0 | 110,739 |
| Zamfara | Anka | 5 | 283 | 143,637 |
| Zamfara | Bakura | 5 | 283 | 187,141 |
| Zamfara | Birnin Magaji | 6 | 339 | 184,083 |
| Zamfara | Bukkuyum | 6 | 339 | 216,348 |
| Zamfara | Bungudu | 8 | 452 | 258,644 |
| Zamfara | Gummi | 17 | 962 | 206,721 |
| Zamfara | Gusau | 19 | 1075 | 383,712 |
| Zamfara | Kaura Namoda | 1 | 57 | 285,363 |
| Zamfara | Maradun | 5 | 283 | 207,563 |
| Zamfara | Maru | 12 | 679 | 293,141 |
| Zamfara | Shinkafi | 3 | 170 | 135,964 |
| Zamfara | Talata Mafara | 16 | 905 | 215,650 |
| Zamfara | Tsafe | 13 | 735 | 266,929 |
| Zamfara | Zurmi | 5 | 283 | 293,977 |
| Total | | 916 | 51,048 | 54,194,698 |

**Supplementary Table 4**

Median smoothed standardized incidence ratio and 95% credible interval indicating noma incidence risk between 1999 and 2024 in 296 LGAs of Northern Nigeria.

| **LGA** | **State** | **Monte Carlo error** | **2.5% interval** | **Median** | **97.5% interval** | **Significant** |
| --- | --- | --- | --- | --- | --- | --- |
| Abadam | Borno | 5.44E-06 | <0.01 | <0.01 | <0.01 | Yes |
| Agaie | Niger | 2.82E-06 | <0.01 | <0.01 | <0.01 | Yes |
| Agwara | Niger | 1.62E-05 | <0.01 | <0.01 | <0.01 | Yes |
| Ajingi | Kano | 5.37E-06 | <0.01 | <0.01 | <0.01 | Yes |
| Albasu | Kano | 4.96E-06 | <0.01 | <0.01 | <0.01 | Yes |
| Aleiro | Kebbi | 0.00265 | 2.19 | 2.57 | 3.00 | Yes |
| Alkaleri | Bauchi | 4.89E-06 | <0.01 | <0.01 | <0.01 | Yes |
| Anka | Zamfara | 0.001824 | 1.86 | 2.09 | 2.34 | Yes |
| Arewa-Dandi | Kebbi | 0.001058 | 0.77 | 0.91 | 1.05 | No |
| Argungu | Kebbi | 7.31E-04 | 0.47 | 0.57 | 0.69 | Yes |
| Askira/Uba | Borno | 2.46E-06 | <0.01 | <0.01 | <0.01 | Yes |
| Augie | Kebbi | 0.002335 | 2.65 | 2.96 | 3.28 | Yes |
| Auyo | Jigawa | 6.33E-06 | <0.01 | <0.01 | <0.01 | Yes |
| Babura | Jigawa | 3.93E-06 | <0.01 | <0.01 | <0.01 | Yes |
| Bade | Yobe | 0.004631 | 11.47 | 12.05 | 12.64 | Yes |
| Bagudo | Kebbi | 9.51E-04 | 1.07 | 1.21 | 1.36 | Yes |
| Bagwai | Kano | 8.16E-06 | <0.01 | <0.01 | <0.01 | Yes |
| Bakori | Katsina | 6.63E-06 | <0.01 | <0.01 | <0.01 | Yes |
| Bakura | Zamfara | 0.001484 | 1.42 | 1.60 | 1.80 | Yes |
| Bama | Borno | 3.26E-06 | <0.01 | <0.01 | <0.01 | Yes |
| Batagarawa | Katsina | 4.19E-06 | <0.01 | <0.01 | <0.01 | Yes |
| Batsari | Katsina | 5.86E-06 | <0.01 | <0.01 | <0.01 | Yes |
| Bauchi | Bauchi | 5.41E-04 | 0.65 | 0.73 | 0.81 | Yes |
| Baure | Katsina | 4.14E-06 | <0.01 | <0.01 | <0.01 | Yes |
| Bayo | Borno | 0.001893 | 1.20 | 1.46 | 1.74 | Yes |
| Bebeji | Kano | 5.77E-04 | 0.29 | 0.38 | 0.47 | Yes |
| Bichi | Kano | 3.14E-06 | <0.01 | <0.01 | <0.01 | Yes |
| Bida | Niger | 4.18E-06 | <0.01 | <0.01 | <0.01 | Yes |
| Bindawa | Katsina | 8.10E-06 | <0.01 | <0.01 | <0.01 | Yes |
| Binji | Sokoto | 0.002581 | 3.19 | 3.56 | 3.95 | Yes |
| Biriniwa | Jigawa | 1.18E-05 | <0.01 | <0.01 | <0.01 | Yes |
| Birnin Kebbi | Kebbi | 0.001935 | 4.23 | 4.49 | 4.75 | Yes |
| Birnin Kudu | Jigawa | 2.33E-06 | <0.01 | <0.01 | <0.01 | Yes |
| Birnin Magaji | Zamfara | 0.001726 | 1.75 | 1.96 | 2.17 | Yes |
| Birnin-Gwari | Kaduna | 5.59E-06 | <0.01 | <0.01 | <0.01 | Yes |
| Biu | Borno | 3.05E-06 | <0.01 | <0.01 | <0.01 | Yes |
| Bodinga | Sokoto | 0.001977 | 3.07 | 3.35 | 3.64 | Yes |
| Bogoro | Bauchi | 1.08E-05 | <0.01 | <0.01 | <0.01 | Yes |
| Borgu | Niger | 8.91E-06 | <0.01 | <0.01 | <0.01 | Yes |
| Bosso | Niger | 4.65E-06 | <0.01 | <0.01 | <0.01 | Yes |
| Buji | Jigawa | 1.22E-05 | <0.01 | <0.01 | <0.01 | Yes |
| Bukkuyum | Zamfara | 0.001289 | 1.50 | 1.66 | 1.85 | Yes |
| Bungudu | Zamfara | 0.001145 | 1.69 | 1.86 | 2.03 | Yes |
| Bunkure | Kano | 7.74E-04 | 0.32 | 0.41 | 0.52 | Yes |
| Bunza | Kebbi | 0.00294 | 4.27 | 4.64 | 5.05 | Yes |
| Bursari | Yobe | 0.002427 | 1.92 | 2.19 | 2.49 | Yes |
| Chanchaga | Niger | 3.58E-06 | <0.01 | <0.01 | <0.01 | Yes |
| Charanchi | Katsina | 8.57E-06 | <0.01 | <0.01 | <0.01 | Yes |
| Chibok | Borno | 1.01E-05 | <0.01 | <0.01 | <0.01 | Yes |
| Chikun | Kaduna | 2.53E-06 | <0.01 | <0.01 | <0.01 | Yes |
| Dala | Kano | 1.86E-06 | <0.01 | <0.01 | <0.01 | Yes |
| Damaturu | Yobe | 0.001503 | 0.72 | 0.91 | 1.13 | No |
| Damban | Bauchi | 0.00184 | 2.16 | 2.40 | 2.66 | Yes |
| Dambatta | Kano | 5.47E-04 | 0.27 | 0.34 | 0.43 | Yes |
| Damboa | Borno | 2.51E-06 | <0.01 | <0.01 | <0.01 | Yes |
| Dan Musa | Katsina | 1.00E-05 | <0.01 | <0.01 | <0.01 | Yes |
| Dandi | Kebbi | 0.001791 | 2.10 | 2.35 | 2.62 | Yes |
| Dandume | Katsina | 0.002643 | 4.28 | 4.65 | 5.05 | Yes |
| Dange Shuni | Sokoto | 0.002385 | 3.83 | 4.12 | 4.43 | Yes |
| Danja | Katsina | 7.94E-06 | <0.01 | <0.01 | <0.01 | Yes |
| Darazo | Bauchi | 5.40E-06 | <0.01 | <0.01 | <0.01 | Yes |
| Dass | Bauchi | 1.52E-05 | <0.01 | <0.01 | <0.01 | Yes |
| Daura | Katsina | 5.26E-06 | <0.01 | <0.01 | <0.01 | Yes |
| Dawakin Kudu | Kano | 6.80E-06 | <0.01 | <0.01 | <0.01 | Yes |
| Dawakin Tofa | Kano | 3.75E-06 | <0.01 | <0.01 | <0.01 | Yes |
| Demsa | Adamawa | 4.57E-06 | <0.01 | <0.01 | <0.01 | Yes |
| Dikwa | Borno | 1.80E-06 | <0.01 | <0.01 | <0.01 | Yes |
| Doguwa | Kano | 9.87E-04 | 0.37 | 0.48 | 0.60 | Yes |
| Dutse | Jigawa | 3.32E-06 | <0.01 | <0.01 | <0.01 | Yes |
| Dutsi | Katsina | 8.15E-06 | <0.01 | <0.01 | <0.01 | Yes |
| Dutsin-Ma | Katsina | 6.44E-06 | <0.01 | <0.01 | <0.01 | Yes |
| Edati | Niger | 4.12E-06 | <0.01 | <0.01 | <0.01 | Yes |
| Fagge | Kano | 3.79E-06 | <0.01 | <0.01 | <0.01 | Yes |
| Fakai | Kebbi | 0.001344 | 0.79 | 0.96 | 1.15 | No |
| Faskari | Katsina | 6.16E-04 | 0.25 | 0.33 | 0.42 | Yes |
| Fika | Yobe | 5.81E-06 | <0.01 | <0.01 | <0.01 | Yes |
| Fufore | Adamawa | 3.41E-06 | <0.01 | <0.01 | <0.01 | Yes |
| Fune | Yobe | 5.49E-06 | <0.01 | <0.01 | <0.01 | Yes |
| Funtua | Katsina | 4.91E-06 | <0.01 | <0.01 | <0.01 | Yes |
| Gabasawa | Kano | 5.29E-06 | <0.01 | <0.01 | <0.01 | Yes |
| Gada | Sokoto | 0.003019 | 10.66 | 11.08 | 11.51 | Yes |
| Gagarawa | Jigawa | 1.06E-05 | <0.01 | <0.01 | <0.01 | Yes |
| Gamawa | Bauchi | 4.06E-06 | <0.01 | <0.01 | <0.01 | Yes |
| Ganjuwa | Bauchi | 4.51E-06 | <0.01 | <0.01 | <0.01 | Yes |
| Ganye | Adamawa | 1.71E-06 | <0.01 | <0.01 | <0.01 | Yes |
| Garki | Jigawa | 6.45E-06 | <0.01 | <0.01 | <0.01 | Yes |
| Garko | Kano | 8.24E-04 | 0.35 | 0.44 | 0.56 | Yes |
| Garum Mallam | Kano | 1.02E-05 | <0.01 | <0.01 | <0.01 | Yes |
| Gaya | Kano | 2.37E-06 | <0.01 | <0.01 | <0.01 | Yes |
| Gbako | Niger | 4.18E-06 | <0.01 | <0.01 | <0.01 | Yes |
| Geidam | Yobe | 6.31E-06 | <0.01 | <0.01 | <0.01 | Yes |
| Gezawa | Kano | 3.19E-06 | <0.01 | <0.01 | <0.01 | Yes |
| Giade | Bauchi | 8.18E-06 | <0.01 | <0.01 | <0.01 | Yes |
| Girei | Adamawa | 0.00167 | 1.70 | 1.94 | 2.20 | Yes |
| Giwa | Kaduna | 3.71E-06 | <0.01 | <0.01 | <0.01 | Yes |
| Gombi | Adamawa | 3.88E-06 | <0.01 | <0.01 | <0.01 | Yes |
| Goronyo | Sokoto | 0.003909 | 11.15 | 11.66 | 12.17 | Yes |
| Gubio | Borno | 6.46E-06 | <0.01 | <0.01 | <0.01 | Yes |
| Gudu | Sokoto | 0.001034 | 0.41 | 0.55 | 0.72 | Yes |
| Gujba | Yobe | 5.68E-06 | <0.01 | <0.01 | <0.01 | Yes |
| Gulani | Yobe | 7.54E-06 | <0.01 | <0.01 | <0.01 | Yes |
| Gumel | Jigawa | 7.55E-06 | <0.01 | <0.01 | <0.01 | Yes |
| Gummi | Zamfara | 0.002015 | 4.64 | 4.94 | 5.25 | Yes |
| Gurara | Niger | 6.10E-06 | <0.01 | <0.01 | <0.01 | Yes |
| Guri | Jigawa | 0.003181 | 6.45 | 6.94 | 7.45 | Yes |
| Gusau | Zamfara | 0.001317 | 2.80 | 2.97 | 3.15 | Yes |
| Guyuk | Adamawa | 5.01E-06 | <0.01 | <0.01 | <0.01 | Yes |
| Guzamala | Borno | 8.20E-06 | <0.01 | <0.01 | <0.01 | Yes |
| Gwadabawa | Sokoto | 0.002587 | 8.33 | 8.71 | 9.11 | Yes |
| Gwale | Kano | 3.78E-06 | <0.01 | <0.01 | <0.01 | Yes |
| Gwandu | Kebbi | 0.001422 | 1.33 | 1.52 | 1.73 | Yes |
| Gwaram | Jigawa | 2.77E-06 | <0.01 | <0.01 | <0.01 | Yes |
| Gwarzo | Kano | 6.95E-04 | 0.31 | 0.39 | 0.50 | Yes |
| Gwiwa | Jigawa | 8.18E-06 | <0.01 | <0.01 | <0.01 | Yes |
| Gwoza | Borno | 1.03E-06 | <0.01 | <0.01 | <0.01 | Yes |
| Hadejia | Jigawa | 7.98E-06 | <0.01 | <0.01 | <0.01 | Yes |
| Hawul | Borno | 3.26E-06 | <0.01 | <0.01 | <0.01 | Yes |
| Hong | Adamawa | 2.08E-06 | <0.01 | <0.01 | <0.01 | Yes |
| Igabi | Kaduna | 8.69E-04 | 1.57 | 1.69 | 1.82 | Yes |
| Ikara | Kaduna | 6.46E-06 | <0.01 | <0.01 | <0.01 | Yes |
| Illela | Sokoto | 0.005245 | 16.64 | 17.33 | 18.04 | Yes |
| Ingawa | Katsina | 6.80E-06 | <0.01 | <0.01 | <0.01 | Yes |
| Isa | Sokoto | 0.003093 | 6.63 | 7.07 | 7.52 | Yes |
| Itas/Gadau | Bauchi | 4.81E-06 | <0.01 | <0.01 | <0.01 | Yes |
| Jaba | Kaduna | 5.30E-06 | <0.01 | <0.01 | <0.01 | Yes |
| Jada | Adamawa | 3.91E-06 | <0.01 | <0.01 | <0.01 | Yes |
| Jahun | Jigawa | 5.67E-04 | 0.27 | 0.34 | 0.42 | Yes |
| Jakusko | Yobe | 6.06E-06 | <0.01 | <0.01 | <0.01 | Yes |
| Jama'are | Bauchi | 8.34E-06 | <0.01 | <0.01 | <0.01 | Yes |
| Jega | Kebbi | 0.001041 | 0.74 | 0.87 | 1.01 | No |
| Jema'a | Kaduna | 2.82E-06 | <0.01 | <0.01 | <0.01 | Yes |
| Jere | Borno | 0.001643 | 2.54 | 2.76 | 3.00 | Yes |
| Jibia | Katsina | 8.11E-04 | 0.30 | 0.39 | 0.49 | Yes |
| Kabo | Kano | 0.001868 | 2.12 | 2.36 | 2.63 | Yes |
| Kachia | Kaduna | 2.20E-06 | <0.01 | <0.01 | <0.01 | Yes |
| Kaduna North | Kaduna | 5.89E-04 | 0.52 | 0.60 | 0.68 | Yes |
| Kaduna South | Kaduna | 4.12E-04 | 0.30 | 0.36 | 0.43 | Yes |
| Kafin Hausa | Jigawa | 4.73E-04 | 0.23 | 0.29 | 0.37 | Yes |
| Kafur | Katsina | 4.53E-06 | <0.01 | <0.01 | <0.01 | Yes |
| Kaga | Borno | 9.70E-06 | <0.01 | <0.01 | <0.01 | Yes |
| Kagarko | Kaduna | 3.55E-06 | <0.01 | <0.01 | <0.01 | Yes |
| Kaita | Katsina | 7.63E-06 | <0.01 | <0.01 | <0.01 | Yes |
| Kajuru | Kaduna | 7.02E-06 | <0.01 | <0.01 | <0.01 | Yes |
| Kala/Balge | Borno | 1.70E-05 | <0.01 | <0.01 | <0.01 | Yes |
| Kalgo | Kebbi | 0.00322 | 2.34 | 2.70 | 3.07 | Yes |
| Kankara | Katsina | 6.66E-06 | <0.01 | <0.01 | <0.01 | Yes |
| Kankia | Katsina | 7.03E-06 | <0.01 | <0.01 | <0.01 | Yes |
| Kano Municipal | Kano | 6.10E-04 | 0.69 | 0.78 | 0.87 | Yes |
| Karasuwa | Yobe | 9.21E-06 | <0.01 | <0.01 | <0.01 | Yes |
| Karaye | Kano | 8.45E-06 | <0.01 | <0.01 | <0.01 | Yes |
| Katagum | Bauchi | 7.47E-04 | 0.82 | 0.92 | 1.04 | No |
| Katcha | Niger | 2.18E-06 | <0.01 | <0.01 | <0.01 | Yes |
| Katsina | Katsina | 6.16E-04 | 0.53 | 0.61 | 0.70 | Yes |
| Kaugama | Jigawa | 4.97E-06 | <0.01 | <0.01 | <0.01 | Yes |
| Kaura | Kaduna | 3.29E-06 | <0.01 | <0.01 | <0.01 | Yes |
| Kaura Namoda | Zamfara | 4.24E-04 | 0.16 | 0.21 | 0.27 | Yes |
| Kauru | Kaduna | 4.33E-06 | <0.01 | <0.01 | <0.01 | Yes |
| Kazaure | Jigawa | 5.30E-06 | <0.01 | <0.01 | <0.01 | Yes |
| Kebbe | Sokoto | 9.01E-04 | 0.32 | 0.43 | 0.56 | Yes |
| Kibiya | Kano | 1.02E-05 | <0.01 | <0.01 | <0.01 | Yes |
| Kirfi | Bauchi | 9.30E-04 | 0.49 | 0.62 | 0.76 | Yes |
| Kiri Kasama | Jigawa | 5.93E-06 | <0.01 | <0.01 | <0.01 | Yes |
| Kiru | Kano | 0.001321 | 2.25 | 2.44 | 2.64 | Yes |
| Kiyawa | Jigawa | 7.55E-06 | <0.01 | <0.01 | <0.01 | Yes |
| Koko/Besse | Kebbi | 0.003141 | 5.19 | 5.56 | 5.95 | Yes |
| Konduga | Borno | 5.11E-06 | <0.01 | <0.01 | <0.01 | Yes |
| Kontagora | Niger | 0.001144 | 0.78 | 0.93 | 1.09 | No |
| Kubau | Kaduna | 5.17E-06 | <0.01 | <0.01 | <0.01 | Yes |
| Kudan | Kaduna | 0.001426 | 0.88 | 1.05 | 1.23 | No |
| Kukawa | Borno | 2.70E-06 | <0.01 | <0.01 | <0.01 | Yes |
| Kumbotso | Kano | 0.001238 | 2.28 | 2.46 | 2.65 | Yes |
| Kunchi | Kano | 7.81E-06 | <0.01 | <0.01 | <0.01 | Yes |
| Kura | Kano | 8.32E-06 | <0.01 | <0.01 | <0.01 | Yes |
| Kurfi | Katsina | 1.08E-05 | <0.01 | <0.01 | <0.01 | Yes |
| Kusada | Katsina | 8.70E-06 | <0.01 | <0.01 | <0.01 | Yes |
| Kware | Sokoto | 0.003052 | 5.15 | 5.54 | 5.97 | Yes |
| Kwaya Kusar | Borno | 1.41E-05 | <0.01 | <0.01 | <0.01 | Yes |
| Lamurde | Adamawa | 6.46E-06 | <0.01 | <0.01 | <0.01 | Yes |
| Lapai | Niger | 6.64E-06 | <0.01 | <0.01 | <0.01 | Yes |
| Lavun | Niger | 4.73E-06 | <0.01 | <0.01 | <0.01 | Yes |
| Lere | Kaduna | 4.63E-06 | <0.01 | <0.01 | <0.01 | Yes |
| Machina | Yobe | 0.00194 | 1.03 | 1.31 | 1.63 | Yes |
| Madagali | Adamawa | 3.54E-06 | <0.01 | <0.01 | <0.01 | Yes |
| Madobi | Kano | 0.001352 | 0.89 | 1.05 | 1.24 | No |
| Mafa | Borno | 6.60E-06 | <0.01 | <0.01 | <0.01 | Yes |
| Magama | Niger | 6.08E-04 | 0.30 | 0.39 | 0.49 | Yes |
| Magumeri | Borno | 4.49E-06 | <0.01 | <0.01 | <0.01 | Yes |
| Mai'adua | Katsina | 3.10E-06 | <0.01 | <0.01 | <0.01 | Yes |
| Maiduguri | Borno | 1.88E-06 | <0.01 | <0.01 | <0.01 | Yes |
| Maigatari | Jigawa | 4.05E-06 | <0.01 | <0.01 | <0.01 | Yes |
| Maiha | Adamawa | 4.69E-06 | <0.01 | <0.01 | <0.01 | Yes |
| Maiyama | Kebbi | 5.82E-04 | 0.25 | 0.33 | 0.42 | Yes |
| Makarfi | Kaduna | 9.91E-06 | <0.01 | <0.01 | <0.01 | Yes |
| Makoda | Kano | 4.01E-06 | <0.01 | <0.01 | <0.01 | Yes |
| Malam Madori | Jigawa | 1.03E-05 | <0.01 | <0.01 | <0.01 | Yes |
| Malumfashi | Katsina | 0.001221 | 1.25 | 1.42 | 1.60 | Yes |
| Mani | Katsina | 6.61E-06 | <0.01 | <0.01 | <0.01 | Yes |
| Maradun | Zamfara | 0.001268 | 1.28 | 1.45 | 1.62 | Yes |
| Mariga | Niger | 6.57E-06 | <0.01 | <0.01 | <0.01 | Yes |
| Marte | Borno | 7.51E-06 | <0.01 | <0.01 | <0.01 | Yes |
| Maru | Zamfara | 0.001292 | 2.28 | 2.46 | 2.64 | Yes |
| Mashegu | Niger | 5.42E-06 | <0.01 | <0.01 | <0.01 | Yes |
| Mashi | Katsina | 9.49E-04 | 0.63 | 0.75 | 0.90 | Yes |
| Matazu | Katsina | 8.73E-06 | <0.01 | <0.01 | <0.01 | Yes |
| Mayo-Belwa | Adamawa | 4.24E-06 | <0.01 | <0.01 | <0.01 | Yes |
| Michika | Adamawa | 3.44E-06 | <0.01 | <0.01 | <0.01 | Yes |
| Miga | Jigawa | 7.73E-06 | <0.01 | <0.01 | <0.01 | Yes |
| Minjibir | Kano | 0.001022 | 0.86 | 0.99 | 1.13 | No |
| Misau | Bauchi | 0.001172 | 1.24 | 1.38 | 1.53 | Yes |
| Mobbar | Borno | 3.65E-06 | <0.01 | <0.01 | <0.01 | Yes |
| Mokwa | Niger | 2.23E-06 | <0.01 | <0.01 | <0.01 | Yes |
| Monguno | Borno | 2.92E-06 | <0.01 | <0.01 | <0.01 | Yes |
| Mubi North | Adamawa | 4.30E-06 | <0.01 | <0.01 | <0.01 | Yes |
| Mubi South | Adamawa | 4.65E-06 | <0.01 | <0.01 | <0.01 | Yes |
| Musawa | Katsina | 4.75E-06 | <0.01 | <0.01 | <0.01 | Yes |
| Muya | Niger | 9.93E-06 | <0.01 | <0.01 | <0.01 | Yes |
| Nangere | Yobe | 1.60E-05 | <0.01 | <0.01 | <0.01 | Yes |
| Nasarawa | Kano | 4.36E-04 | 0.43 | 0.49 | 0.54 | Yes |
| Ngala | Borno | 4.09E-06 | <0.01 | <0.01 | <0.01 | Yes |
| Nganzai | Borno | 1.39E-05 | <0.01 | <0.01 | <0.01 | Yes |
| Ngaski | Kebbi | 7.72E-06 | <0.01 | <0.01 | <0.01 | Yes |
| Nguru | Yobe | 8.57E-04 | 0.42 | 0.53 | 0.66 | Yes |
| Ningi | Bauchi | 3.95E-04 | 0.19 | 0.23 | 0.28 | Yes |
| Numan | Adamawa | 4.01E-06 | <0.01 | <0.01 | <0.01 | Yes |
| Paikoro | Niger | 2.69E-06 | <0.01 | <0.01 | <0.01 | Yes |
| Potiskum | Yobe | 3.56E-06 | <0.01 | <0.01 | <0.01 | Yes |
| Rabah | Sokoto | 0.002809 | 5.65 | 6.04 | 6.47 | Yes |
| Rafi | Niger | 5.90E-06 | <0.01 | <0.01 | <0.01 | Yes |
| Rano | Kano | 0.002079 | 2.19 | 2.44 | 2.72 | Yes |
| Rijau | Niger | 7.07E-04 | 0.31 | 0.40 | 0.50 | Yes |
| Rimi | Katsina | 0.001623 | 1.48 | 1.68 | 1.90 | Yes |
| Rimin Gado | Kano | 1.03E-05 | <0.01 | <0.01 | <0.01 | Yes |
| Ringim | Jigawa | 3.60E-06 | <0.01 | <0.01 | <0.01 | Yes |
| Rogo | Kano | 6.70E-06 | <0.01 | <0.01 | <0.01 | Yes |
| Roni | Jigawa | 8.10E-06 | <0.01 | <0.01 | <0.01 | Yes |
| Sabon Birni | Sokoto | 0.003202 | 9.29 | 9.72 | 10.16 | Yes |
| Sabon-Gari | Kaduna | 3.72E-06 | <0.01 | <0.01 | <0.01 | Yes |
| Sabuwa | Katsina | 5.74E-06 | <0.01 | <0.01 | <0.01 | Yes |
| Safana | Katsina | 5.68E-04 | 0.27 | 0.35 | 0.44 | Yes |
| Sakaba | Kebbi | 1.31E-05 | <0.01 | <0.01 | <0.01 | Yes |
| Sandamu | Katsina | 8.81E-04 | 0.37 | 0.47 | 0.60 | Yes |
| Sanga | Kaduna | 2.63E-06 | <0.01 | <0.01 | <0.01 | Yes |
| Shagari | Sokoto | 0.003257 | 6.35 | 6.76 | 7.19 | Yes |
| Shanga | Kebbi | 9.60E-04 | 0.34 | 0.45 | 0.58 | Yes |
| Shani | Borno | 6.40E-06 | <0.01 | <0.01 | <0.01 | Yes |
| Shanono | Kano | 1.27E-05 | <0.01 | <0.01 | <0.01 | Yes |
| Shelleng | Adamawa | 2.76E-06 | <0.01 | <0.01 | <0.01 | Yes |
| Shinkafi | Zamfara | 0.001517 | 1.13 | 1.33 | 1.53 | Yes |
| Shira | Bauchi | 3.67E-06 | <0.01 | <0.01 | <0.01 | Yes |
| Shiroro | Niger | 0.001225 | 1.34 | 1.50 | 1.67 | Yes |
| Silame | Sokoto | 0.002345 | 2.72 | 3.04 | 3.40 | Yes |
| Soba | Kaduna | 4.21E-06 | <0.01 | <0.01 | <0.01 | Yes |
| Sokoto North | Sokoto | 0.003042 | 10.52 | 10.93 | 11.39 | Yes |
| Sokoto South | Sokoto | 0.002112 | 4.01 | 4.29 | 4.60 | Yes |
| Song | Adamawa | 4.50E-06 | <0.01 | <0.01 | <0.01 | Yes |
| Sule Tankarkar | Jigawa | 4.95E-06 | <0.01 | <0.01 | <0.01 | Yes |
| Suleja | Niger | 3.77E-06 | <0.01 | <0.01 | <0.01 | Yes |
| Sumaila | Kano | 3.83E-06 | <0.01 | <0.01 | <0.01 | Yes |
| Suru | Kebbi | 0.001487 | 2.07 | 2.32 | 2.58 | Yes |
| Tafa | Niger | 7.42E-06 | <0.01 | <0.01 | <0.01 | Yes |
| Tafawa-Balewa | Bauchi | 2.42E-06 | <0.01 | <0.01 | <0.01 | Yes |
| Takai | Kano | 6.54E-06 | <0.01 | <0.01 | <0.01 | Yes |
| Talata Mafara | Zamfara | 0.002219 | 4.18 | 4.45 | 4.75 | Yes |
| Tambuwal | Sokoto | 0.002538 | 5.08 | 5.40 | 5.72 | Yes |
| Tangaza | Sokoto | 0.004032 | 10.47 | 11.09 | 11.75 | Yes |
| Tarauni | Kano | 6.22E-06 | <0.01 | <0.01 | <0.01 | Yes |
| Tarmua | Yobe | 1.20E-05 | <0.01 | <0.01 | <0.01 | Yes |
| Taura | Jigawa | 6.77E-06 | <0.01 | <0.01 | <0.01 | Yes |
| Tofa | Kano | 1.18E-05 | <0.01 | <0.01 | <0.01 | Yes |
| Toro | Bauchi | 3.11E-06 | <0.01 | <0.01 | <0.01 | Yes |
| Toungo | Adamawa | 8.63E-06 | <0.01 | <0.01 | <0.01 | Yes |
| Tsafe | Zamfara | 0.001589 | 2.72 | 2.92 | 3.13 | Yes |
| Tsanyawa | Kano | 5.33E-06 | <0.01 | <0.01 | <0.01 | Yes |
| Tudun Wada | Kano | 6.25E-06 | <0.01 | <0.01 | <0.01 | Yes |
| Tureta | Sokoto | 0.003268 | 3.41 | 3.88 | 4.39 | Yes |
| Ungogo | Kano | 9.07E-04 | 1.26 | 1.39 | 1.52 | Yes |
| Wamako | Sokoto | 0.0038 | 13.36 | 13.92 | 14.48 | Yes |
| Warawa | Kano | 7.63E-06 | <0.01 | <0.01 | <0.01 | Yes |
| Warji | Bauchi | 1.26E-05 | <0.01 | <0.01 | <0.01 | Yes |
| Wasagu/Danko | Kebbi | 9.45E-04 | 1.36 | 1.52 | 1.67 | Yes |
| Wudil | Kano | 4.30E-06 | <0.01 | <0.01 | <0.01 | Yes |
| Wurno | Sokoto | 0.001235 | 0.83 | 0.98 | 1.15 | No |
| Wushishi | Niger | 6.06E-06 | <0.01 | <0.01 | <0.01 | Yes |
| Yabo | Sokoto | 0.002824 | 4.65 | 5.06 | 5.51 | Yes |
| Yankwashi | Jigawa | 7.36E-06 | <0.01 | <0.01 | <0.01 | Yes |
| Yauri | Kebbi | 0.002271 | 1.99 | 2.28 | 2.59 | Yes |
| Yola North | Adamawa | 7.07E-04 | 0.52 | 0.63 | 0.75 | Yes |
| Yola South | Adamawa | 5.11E-06 | <0.01 | <0.01 | <0.01 | Yes |
| Yunusari | Yobe | 7.75E-06 | <0.01 | <0.01 | <0.01 | Yes |
| Yusufari | Yobe | 9.81E-06 | <0.01 | <0.01 | <0.01 | Yes |
| Zaki | Bauchi | 6.91E-04 | 0.38 | 0.47 | 0.58 | Yes |
| Zango | Katsina | 5.10E-06 | <0.01 | <0.01 | <0.01 | Yes |
| Zangon-Kataf | Kaduna | 2.64E-06 | <0.01 | <0.01 | <0.01 | Yes |
| Zaria | Kaduna | 6.18E-04 | 0.63 | 0.71 | 0.80 | Yes |
| Zurmi | Zamfara | 8.52E-04 | 0.91 | 1.02 | 1.14 | No |
| Zuru | Kebbi | 0.001796 | 2.86 | 3.13 | 3.41 | Yes |

**Supplementary Table 5**

Stratified noma incidence risk estimates for different LGAs by sex

| **State** | **LGA** | **Median SIR**  **(Females)** | **Significant**  **(Females)** | **Median SIR (Males)** | **Significant (Males)** |
| --- | --- | --- | --- | --- | --- |
| Borno | Abadam | <0.01 | Yes | <0.01 | Yes |
| Niger | Agaie | <0.01 | Yes | <0.01 | Yes |
| Niger | Agwara | <0.01 | Yes | <0.01 | Yes |
| Kano | Ajingi | <0.01 | Yes | <0.01 | Yes |
| Kano | Albasu | <0.01 | Yes | <0.01 | Yes |
| Kebbi | Aleiro | 3.56 | Yes | 1.61 | Yes |
| Bauchi | Alkaleri | <0.01 | Yes | <0.01 | Yes |
| Zamfara | Anka | <0.01 | Yes | 4.00 | Yes |
| Kebbi | Arewa-Dandi | <0.01 | Yes | 1.75 | Yes |
| Kebbi | Argungu | 0.59 | Yes | 0.55 | Yes |
| Borno | Askira/Uba | <0.01 | Yes | <0.01 | Yes |
| Kebbi | Augie | 1.03 | No | 4.73 | Yes |
| Jigawa | Auyo | <0.01 | Yes | <0.01 | Yes |
| Jigawa | Babura | <0.01 | Yes | <0.01 | Yes |
| Yobe | Bade | 17.44 | Yes | 7.43 | Yes |
| Kebbi | Bagudo | 1.51 | Yes | 0.92 | No |
| Kano | Bagwai | <0.01 | Yes | <0.01 | Yes |
| Katsina | Bakori | <0.01 | Yes | <0.01 | Yes |
| Zamfara | Bakura | 0.67 | Yes | 2.47 | Yes |
| Borno | Bama | <0.01 | Yes | <0.01 | Yes |
| Katsina | Batagarawa | <0.01 | Yes | <0.01 | Yes |
| Katsina | Batsari | <0.01 | Yes | <0.01 | Yes |
| Bauchi | Bauchi | <0.01 | Yes | 1.39 | Yes |
| Katsina | Baure | <0.01 | Yes | <0.01 | Yes |
| Borno | Bayo | <0.01 | Yes | 2.69 | Yes |
| Kano | Bebeji | 0.83 | No | <0.01 | Yes |
| Kano | Bichi | <0.01 | Yes | <0.01 | Yes |
| Niger | Bida | <0.01 | Yes | <0.01 | Yes |
| Katsina | Bindawa | <0.01 | Yes | <0.01 | Yes |
| Sokoto | Binji | 3.16 | Yes | 3.91 | Yes |
| Jigawa | Biriniwa | <0.01 | Yes | <0.01 | Yes |
| Kebbi | Birnin Kebbi | 1.79 | Yes | 6.96 | Yes |
| Jigawa | Birnin Kudu | <0.01 | Yes | <0.01 | Yes |
| Zamfara | Birnin Magaji | 2.74 | Yes | 1.26 | Yes |
| Kaduna | Birnin-Gwari | <0.01 | Yes | <0.01 | Yes |
| Borno | Biu | <0.01 | Yes | <0.01 | Yes |
| Sokoto | Bodinga | 1.27 | Yes | 5.23 | Yes |
| Bauchi | Bogoro | <0.01 | Yes | <0.01 | Yes |
| Niger | Borgu | <0.01 | Yes | <0.01 | Yes |
| Niger | Bosso | <0.01 | Yes | <0.01 | Yes |
| Jigawa | Buji | <0.01 | Yes | <0.01 | Yes |
| Zamfara | Bukkuyum | 1.74 | Yes | 1.59 | Yes |
| Zamfara | Bungudu | 1.45 | Yes | 2.24 | Yes |
| Kano | Bunkure | 0.90 | No | <0.01 | Yes |
| Kebbi | Bunza | 5.79 | Yes | 3.55 | Yes |
| Yobe | Bursari | 4.78 | Yes | <0.01 | Yes |
| Niger | Chanchaga | <0.01 | Yes | <0.01 | Yes |
| Katsina | Charanchi | <0.01 | Yes | <0.01 | Yes |
| Borno | Chibok | <0.01 | Yes | <0.01 | Yes |
| Kaduna | Chikun | <0.01 | Yes | <0.01 | Yes |
| Kano | Dala | <0.01 | Yes | <0.01 | Yes |
| Yobe | Damaturu | <0.01 | Yes | 1.67 | Yes |
| Bauchi | Damban | 3.84 | Yes | 1.14 | No |
| Kano | Dambatta | 0.76 | Yes | <0.01 | Yes |
| Borno | Damboa | <0.01 | Yes | <0.01 | Yes |
| Katsina | Dan Musa | <0.01 | Yes | <0.01 | Yes |
| Kebbi | Dandi | 1.63 | Yes | 3.01 | Yes |
| Katsina | Dandume | <0.01 | Yes | 7.57 | Yes |
| Sokoto | Dange Shuni | 4.63 | Yes | 3.67 | Yes |
| Katsina | Danja | <0.01 | Yes | <0.01 | Yes |
| Bauchi | Darazo | <0.01 | Yes | <0.01 | Yes |
| Bauchi | Dass | <0.01 | Yes | <0.01 | Yes |
| Katsina | Daura | <0.01 | Yes | <0.01 | Yes |
| Kano | Dawakin Kudu | <0.01 | Yes | <0.01 | Yes |
| Kano | Dawakin Tofa | <0.01 | Yes | <0.01 | Yes |
| Adamawa | Demsa | <0.01 | Yes | <0.01 | Yes |
| Borno | Dikwa | <0.01 | Yes | <0.01 | Yes |
| Kano | Doguwa | 1.06 | No | <0.01 | Yes |
| Jigawa | Dutse | <0.01 | Yes | <0.01 | Yes |
| Katsina | Dutsi | <0.01 | Yes | <0.01 | Yes |
| Katsina | Dutsin-Ma | <0.01 | Yes | <0.01 | Yes |
| Niger | Edati | <0.01 | Yes | <0.01 | Yes |
| Kano | Fagge | <0.01 | Yes | <0.01 | Yes |
| Kebbi | Fakai | 1.99 | Yes | <0.01 | Yes |
| Katsina | Faskari | 0.71 | Yes | <0.01 | Yes |
| Yobe | Fika | <0.01 | Yes | <0.01 | Yes |
| Adamawa | Fufore | <0.01 | Yes | <0.01 | Yes |
| Yobe | Fune | <0.01 | Yes | <0.01 | Yes |
| Katsina | Funtua | <0.01 | Yes | <0.01 | Yes |
| Kano | Gabasawa | <0.01 | Yes | <0.01 | Yes |
| Sokoto | Gada | 13.43 | Yes | 8.97 | Yes |
| Jigawa | Gagarawa | <0.01 | Yes | <0.01 | Yes |
| Bauchi | Gamawa | <0.01 | Yes | <0.01 | Yes |
| Bauchi | Ganjuwa | <0.01 | Yes | <0.01 | Yes |
| Adamawa | Ganye | <0.01 | Yes | <0.01 | Yes |
| Jigawa | Garki | <0.01 | Yes | <0.01 | Yes |
| Kano | Garko | 0.98 | No | <0.01 | Yes |
| Kano | Garum Mallam | <0.01 | Yes | <0.01 | Yes |
| Kano | Gaya | <0.01 | Yes | <0.01 | Yes |
| Niger | Gbako | <0.01 | Yes | <0.01 | Yes |
| Yobe | Geidam | <0.01 | Yes | <0.01 | Yes |
| Kano | Gezawa | <0.01 | Yes | <0.01 | Yes |
| Bauchi | Giade | <0.01 | Yes | <0.01 | Yes |
| Adamawa | Girei | <0.01 | Yes | 3.69 | Yes |
| Kaduna | Giwa | <0.01 | Yes | <0.01 | Yes |
| Adamawa | Gombi | <0.01 | Yes | <0.01 | Yes |
| Sokoto | Goronyo | 6.12 | Yes | 16.72 | Yes |
| Borno | Gubio | <0.01 | Yes | <0.01 | Yes |
| Sokoto | Gudu | <0.01 | Yes | 1.03 | No |
| Yobe | Gujba | <0.01 | Yes | <0.01 | Yes |
| Yobe | Gulani | <0.01 | Yes | <0.01 | Yes |
| Jigawa | Gumel | <0.01 | Yes | <0.01 | Yes |
| Zamfara | Gummi | 3.04 | Yes | 6.69 | Yes |
| Niger | Gurara | <0.01 | Yes | <0.01 | Yes |
| Jigawa | Guri | 14.62 | Yes | <0.01 | Yes |
| Zamfara | Gusau | 2.30 | Yes | 3.60 | Yes |
| Adamawa | Guyuk | <0.01 | Yes | <0.01 | Yes |
| Borno | Guzamala | <0.01 | Yes | <0.01 | Yes |
| Sokoto | Gwadabawa | 9.17 | Yes | 8.32 | Yes |
| Kano | Gwale | <0.01 | Yes | <0.01 | Yes |
| Kebbi | Gwandu | 0.79 | No | 2.21 | Yes |
| Jigawa | Gwaram | <0.01 | Yes | <0.01 | Yes |
| Kano | Gwarzo | 0.87 | No | <0.01 | Yes |
| Jigawa | Gwiwa | <0.01 | Yes | <0.01 | Yes |
| Borno | Gwoza | <0.01 | Yes | <0.01 | Yes |
| Jigawa | Hadejia | <0.01 | Yes | <0.01 | Yes |
| Borno | Hawul | <0.01 | Yes | <0.01 | Yes |
| Adamawa | Hong | <0.01 | Yes | <0.01 | Yes |
| Kaduna | Igabi | 3.56 | Yes | <0.01 | Yes |
| Kaduna | Ikara | <0.01 | Yes | <0.01 | Yes |
| Sokoto | Illela | 14.07 | Yes | 20.30 | Yes |
| Katsina | Ingawa | <0.01 | Yes | <0.01 | Yes |
| Sokoto | Isa | 5.97 | Yes | 8.11 | Yes |
| Bauchi | Itas/Gadau | <0.01 | Yes | <0.01 | Yes |
| Kaduna | Jaba | <0.01 | Yes | <0.01 | Yes |
| Adamawa | Jada | <0.01 | Yes | <0.01 | Yes |
| Jigawa | Jahun | 0.71 | Yes | <0.01 | Yes |
| Yobe | Jakusko | <0.01 | Yes | <0.01 | Yes |
| Bauchi | Jama'are | <0.01 | Yes | <0.01 | Yes |
| Kebbi | Jega | <0.01 | Yes | 1.68 | Yes |
| Kaduna | Jema'a | <0.01 | Yes | <0.01 | Yes |
| Borno | Jere | 5.97 | Yes | <0.01 | Yes |
| Katsina | Jibia | 0.81 | No | <0.01 | Yes |
| Kano | Kabo | 1.04 | No | 3.46 | Yes |
| Kaduna | Kachia | <0.01 | Yes | <0.01 | Yes |
| Kaduna | Kaduna North | 0.42 | Yes | 0.76 | Yes |
| Kaduna | Kaduna South | 0.38 | Yes | 0.34 | Yes |
| Jigawa | Kafin Hausa | 0.62 | Yes | <0.01 | Yes |
| Katsina | Kafur | <0.01 | Yes | <0.01 | Yes |
| Borno | Kaga | <0.01 | Yes | <0.01 | Yes |
| Kaduna | Kagarko | <0.01 | Yes | <0.01 | Yes |
| Katsina | Kaita | <0.01 | Yes | <0.01 | Yes |
| Kaduna | Kajuru | <0.01 | Yes | <0.01 | Yes |
| Borno | Kala/Balge | <0.01 | Yes | <0.01 | Yes |
| Kebbi | Kalgo | 5.66 | Yes | <0.01 | Yes |
| Katsina | Kankara | <0.01 | Yes | <0.01 | Yes |
| Katsina | Kankia | <0.01 | Yes | <0.01 | Yes |
| Kano | Kano Municipal | 1.28 | Yes | 0.36 | Yes |
| Yobe | Karasuwa | <0.01 | Yes | <0.01 | Yes |
| Kano | Karaye | <0.01 | Yes | <0.01 | Yes |
| Bauchi | Katagum | 1.96 | Yes | <0.01 | Yes |
| Niger | Katcha | <0.01 | Yes | <0.01 | Yes |
| Katsina | Katsina | 0.86 | No | 0.38 | Yes |
| Jigawa | Kaugama | <0.01 | Yes | <0.01 | Yes |
| Kaduna | Kaura | <0.01 | Yes | <0.01 | Yes |
| Zamfara | Kaura Namoda | 0.44 | Yes | <0.01 | Yes |
| Kaduna | Kauru | <0.01 | Yes | <0.01 | Yes |
| Jigawa | Kazaure | <0.01 | Yes | <0.01 | Yes |
| Sokoto | Kebbe | 0.90 | No | <0.01 | Yes |
| Kano | Kibiya | <0.01 | Yes | <0.01 | Yes |
| Bauchi | Kirfi | <0.01 | Yes | 1.18 | No |
| Jigawa | Kiri Kasama | <0.01 | Yes | <0.01 | Yes |
| Kano | Kiru | 2.96 | Yes | 1.98 | Yes |
| Jigawa | Kiyawa | <0.01 | Yes | <0.01 | Yes |
| Kebbi | Koko/Besse | 6.93 | Yes | 4.26 | Yes |
| Borno | Konduga | <0.01 | Yes | <0.01 | Yes |
| Niger | Kontagora | 0.98 | No | 0.88 | No |
| Kaduna | Kubau | <0.01 | Yes | <0.01 | Yes |
| Kaduna | Kudan | 1.11 | No | 0.99 | No |
| Borno | Kukawa | <0.01 | Yes | <0.01 | Yes |
| Kano | Kumbotso | 3.79 | Yes | 1.35 | Yes |
| Kano | Kunchi | <0.01 | Yes | <0.01 | Yes |
| Kano | Kura | <0.01 | Yes | <0.01 | Yes |
| Katsina | Kurfi | <0.01 | Yes | <0.01 | Yes |
| Katsina | Kusada | <0.01 | Yes | <0.01 | Yes |
| Sokoto | Kware | 6.63 | Yes | 4.53 | Yes |
| Borno | Kwaya Kusar | <0.01 | Yes | <0.01 | Yes |
| Adamawa | Lamurde | <0.01 | Yes | <0.01 | Yes |
| Niger | Lapai | <0.01 | Yes | <0.01 | Yes |
| Niger | Lavun | <0.01 | Yes | <0.01 | Yes |
| Kaduna | Lere | <0.01 | Yes | <0.01 | Yes |
| Yobe | Machina | 2.84 | Yes | <0.01 | Yes |
| Adamawa | Madagali | <0.01 | Yes | <0.01 | Yes |
| Kano | Madobi | 2.31 | Yes | <0.01 | Yes |
| Borno | Mafa | <0.01 | Yes | <0.01 | Yes |
| Niger | Magama | <0.01 | Yes | 0.74 | Yes |
| Borno | Magumeri | <0.01 | Yes | <0.01 | Yes |
| Katsina | Mai'adua | <0.01 | Yes | <0.01 | Yes |
| Borno | Maiduguri | <0.01 | Yes | <0.01 | Yes |
| Jigawa | Maigatari | <0.01 | Yes | <0.01 | Yes |
| Adamawa | Maiha | <0.01 | Yes | <0.01 | Yes |
| Kebbi | Maiyama | <0.01 | Yes | 0.63 | Yes |
| Kaduna | Makarfi | <0.01 | Yes | <0.01 | Yes |
| Kano | Makoda | <0.01 | Yes | <0.01 | Yes |
| Jigawa | Malam Madori | <0.01 | Yes | <0.01 | Yes |
| Katsina | Malumfashi | <0.01 | Yes | 2.69 | Yes |
| Katsina | Mani | <0.01 | Yes | <0.01 | Yes |
| Zamfara | Maradun | 1.82 | Yes | 1.11 | No |
| Niger | Mariga | <0.01 | Yes | <0.01 | Yes |
| Borno | Marte | <0.01 | Yes | <0.01 | Yes |
| Zamfara | Maru | 3.84 | Yes | 1.18 | Yes |
| Niger | Mashegu | <0.01 | Yes | <0.01 | Yes |
| Katsina | Mashi | 1.61 | Yes | <0.01 | Yes |
| Katsina | Matazu | <0.01 | Yes | <0.01 | Yes |
| Adamawa | Mayo-Belwa | <0.01 | Yes | <0.01 | Yes |
| Adamawa | Michika | <0.01 | Yes | <0.01 | Yes |
| Jigawa | Miga | <0.01 | Yes | <0.01 | Yes |
| Kano | Minjibir | 2.18 | Yes | <0.01 | Yes |
| Bauchi | Misau | <0.01 | Yes | 2.59 | Yes |
| Borno | Mobbar | <0.01 | Yes | <0.01 | Yes |
| Niger | Mokwa | <0.01 | Yes | <0.01 | Yes |
| Borno | Monguno | <0.01 | Yes | <0.01 | Yes |
| Adamawa | Mubi North | <0.01 | Yes | <0.01 | Yes |
| Adamawa | Mubi South | <0.01 | Yes | <0.01 | Yes |
| Katsina | Musawa | <0.01 | Yes | <0.01 | Yes |
| Niger | Muya | <0.01 | Yes | <0.01 | Yes |
| Yobe | Nangere | <0.01 | Yes | <0.01 | Yes |
| Kano | Nasarawa | <0.01 | Yes | 0.89 | No |
| Borno | Ngala | <0.01 | Yes | <0.01 | Yes |
| Borno | Nganzai | <0.01 | Yes | <0.01 | Yes |
| Kebbi | Ngaski | <0.01 | Yes | <0.01 | Yes |
| Yobe | Nguru | <0.01 | Yes | 0.98 | No |
| Bauchi | Ningi | 0.49 | Yes | <0.01 | Yes |
| Adamawa | Numan | <0.01 | Yes | <0.01 | Yes |
| Niger | Paikoro | <0.01 | Yes | <0.01 | Yes |
| Yobe | Potiskum | <0.01 | Yes | <0.01 | Yes |
| Sokoto | Rabah | 5.97 | Yes | 6.13 | Yes |
| Niger | Rafi | <0.01 | Yes | <0.01 | Yes |
| Kano | Rano | <0.01 | Yes | 4.49 | Yes |
| Niger | Rijau | <0.01 | Yes | 0.75 | Yes |
| Katsina | Rimi | <0.01 | Yes | 3.19 | Yes |
| Kano | Rimin Gado | <0.01 | Yes | <0.01 | Yes |
| Jigawa | Ringim | <0.01 | Yes | <0.01 | Yes |
| Kano | Rogo | <0.01 | Yes | <0.01 | Yes |
| Jigawa | Roni | <0.01 | Yes | <0.01 | Yes |
| Sokoto | Sabon Birni | 8.07 | Yes | 11.27 | Yes |
| Kaduna | Sabon-Gari | <0.01 | Yes | <0.01 | Yes |
| Katsina | Sabuwa | <0.01 | Yes | <0.01 | Yes |
| Katsina | Safana | 0.73 | Yes | <0.01 | Yes |
| Kebbi | Sakaba | <0.01 | Yes | <0.01 | Yes |
| Katsina | Sandamu | 1.00 | No | <0.01 | Yes |
| Kaduna | Sanga | <0.01 | Yes | <0.01 | Yes |
| Sokoto | Shagari | 7.12 | Yes | 6.45 | Yes |
| Kebbi | Shanga | <0.01 | Yes | 0.86 | No |
| Borno | Shani | <0.01 | Yes | <0.01 | Yes |
| Kano | Shanono | <0.01 | Yes | <0.01 | Yes |
| Adamawa | Shelleng | <0.01 | Yes | <0.01 | Yes |
| Zamfara | Shinkafi | 2.77 | Yes | <0.01 | Yes |
| Bauchi | Shira | <0.01 | Yes | <0.01 | Yes |
| Niger | Shiroro | <0.01 | Yes | 2.84 | Yes |
| Sokoto | Silame | 6.33 | Yes | <0.01 | Yes |
| Kaduna | Soba | <0.01 | Yes | <0.01 | Yes |
| Sokoto | Sokoto North | 12.40 | Yes | 9.60 | Yes |
| Sokoto | Sokoto South | 3.96 | Yes | 4.62 | Yes |
| Adamawa | Song | <0.01 | Yes | <0.01 | Yes |
| Jigawa | Sule Tankarkar | <0.01 | Yes | <0.01 | Yes |
| Niger | Suleja | <0.01 | Yes | <0.01 | Yes |
| Kano | Sumaila | <0.01 | Yes | <0.01 | Yes |
| Kebbi | Suru | 2.43 | Yes | 2.21 | Yes |
| Niger | Tafa | <0.01 | Yes | <0.01 | Yes |
| Bauchi | Tafawa-Balewa | <0.01 | Yes | <0.01 | Yes |
| Kano | Takai | <0.01 | Yes | <0.01 | Yes |
| Zamfara | Talata Mafara | 4.08 | Yes | 4.80 | Yes |
| Sokoto | Tambuwal | 4.44 | Yes | 6.28 | Yes |
| Sokoto | Tangaza | 4.88 | Yes | 16.80 | Yes |
| Kano | Tarauni | <0.01 | Yes | <0.01 | Yes |
| Yobe | Tarmua | <0.01 | Yes | <0.01 | Yes |
| Jigawa | Taura | <0.01 | Yes | <0.01 | Yes |
| Kano | Tofa | <0.01 | Yes | <0.01 | Yes |
| Bauchi | Toro | <0.01 | Yes | <0.01 | Yes |
| Adamawa | Toungo | <0.01 | Yes | <0.01 | Yes |
| Zamfara | Tsafe | 1.88 | Yes | 3.91 | Yes |
| Kano | Tsanyawa | <0.01 | Yes | <0.01 | Yes |
| Kano | Tudun Wada | <0.01 | Yes | <0.01 | Yes |
| Sokoto | Tureta | 3.21 | Yes | 4.42 | Yes |
| Kano | Ungogo | 2.19 | Yes | 0.72 | Yes |
| Sokoto | Wamako | 18.01 | Yes | 10.18 | Yes |
| Kano | Warawa | <0.01 | Yes | <0.01 | Yes |
| Bauchi | Warji | <0.01 | Yes | <0.01 | Yes |
| Kebbi | Wasagu/Danko | 1.34 | Yes | 1.65 | Yes |
| Kano | Wudil | <0.01 | Yes | <0.01 | Yes |
| Sokoto | Wurno | 1.40 | Yes | 0.62 | Yes |
| Niger | Wushishi | <0.01 | Yes | <0.01 | Yes |
| Sokoto | Yabo | 0.96 | No | 8.83 | Yes |
| Jigawa | Yankwashi | <0.01 | Yes | <0.01 | Yes |
| Kebbi | Yauri | 3.54 | Yes | 1.09 | No |
| Adamawa | Yola North | 1.33 | Yes | <0.01 | Yes |
| Adamawa | Yola South | <0.01 | Yes | <0.01 | Yes |
| Yobe | Yunusari | <0.01 | Yes | <0.01 | Yes |
| Yobe | Yusufari | <0.01 | Yes | <0.01 | Yes |
| Bauchi | Zaki | <0.01 | Yes | 0.89 | No |
| Katsina | Zango | <0.01 | Yes | <0.01 | Yes |
| Kaduna | Zangon-Kataf | <0.01 | Yes | <0.01 | Yes |
| Kaduna | Zaria | 1.13 | No | 0.34 | Yes |
| Zamfara | Zurmi | <0.01 | Yes | 1.95 | Yes |
| Kebbi | Zuru | <0.01 | Yes | 6.01 | Yes |

**Supplementary Table 5**

Median SIR estimates indicating noma incidence risk at different time intervals for all LGAs in 12 Northern Nigerian states. (Median SIRs in bold are significantly higher than the average relative risk of noma incidence in Northern Nigeria).

| **States** | **LGA** | **1999-2004** | **2005-2009** | **2010-2014** | **2015-2019** | **2020-2024** |
| --- | --- | --- | --- | --- | --- | --- |
| Adamawa | Demsa | <0.01 | <0.01 | <0.01 | <0.01 | <0.01 |
|  | Fufore | <0.01 | <0.01 | <0.01 | <0.01 | <0.01 |
|  | Ganye | <0.01 | <0.01 | <0.01 | <0.01 | <0.01 |
|  | Girei | <0.01 | <0.01 | <0.01 | <0.01 | **2.81** |
|  | Gombi | <0.01 | <0.01 | <0.01 | <0.01 | <0.01 |
|  | Guyuk | <0.01 | <0.01 | <0.01 | <0.01 | <0.01 |
|  | Hong | <0.01 | <0.01 | <0.01 | <0.01 | <0.01 |
|  | Jada | <0.01 | <0.01 | <0.01 | <0.01 | <0.01 |
|  | Lamurde | <0.01 | <0.01 | <0.01 | <0.01 | <0.01 |
|  | Madagali | <0.01 | <0.01 | <0.01 | <0.01 | <0.01 |
|  | Maiha | <0.01 | <0.01 | <0.01 | <0.01 | <0.01 |
|  | Mayo-Belwa | <0.01 | <0.01 | <0.01 | <0.01 | <0.01 |
|  | Michika | <0.01 | <0.01 | <0.01 | <0.01 | <0.01 |
|  | Mubi North | <0.01 | <0.01 | <0.01 | <0.01 | <0.01 |
|  | Mubi South | <0.01 | <0.01 | <0.01 | <0.01 | <0.01 |
|  | Numan | <0.01 | <0.01 | <0.01 | <0.01 | <0.01 |
|  | Shelleng | <0.01 | <0.01 | <0.01 | <0.01 | <0.01 |
|  | Song | <0.01 | <0.01 | <0.01 | <0.01 | <0.01 |
|  | Toungo | <0.01 | <0.01 | <0.01 | <0.01 | <0.01 |
|  | Yola North | **6.35** | <0.01 | <0.01 | <0.01 | <0.01 |
|  | Yola South | <0.01 | <0.01 | <0.01 | <0.01 | <0.01 |
| Bauchi | Alkaleri | <0.01 | <0.01 | <0.01 | <0.01 | <0.01 |
|  | Bauchi | <0.01 | <0.01 | <0.01 | <0.01 | 1.07 |
|  | Bogoro | <0.01 | <0.01 | <0.01 | <0.01 | <0.01 |
|  | Damban | <0.01 | <0.01 | <0.01 | <0.01 | **3.52** |
|  | Darazo | <0.01 | <0.01 | <0.01 | <0.01 | <0.01 |
|  | Dass | <0.01 | <0.01 | <0.01 | <0.01 | <0.01 |
|  | Gamawa | <0.01 | <0.01 | <0.01 | <0.01 | <0.01 |
|  | Ganjuwa | <0.01 | <0.01 | <0.01 | <0.01 | <0.01 |
|  | Giade | <0.01 | <0.01 | <0.01 | <0.01 | <0.01 |
|  | Itas/Gadau | <0.01 | <0.01 | <0.01 | <0.01 | <0.01 |
|  | Jama'are | <0.01 | <0.01 | <0.01 | <0.01 | <0.01 |
|  | Katagum | <0.01 | <0.01 | <0.01 | **2.42** | 0.90 |
|  | Kirfi | <0.01 | <0.01 | <0.01 | <0.01 | 0.89 |
|  | Misau | <0.01 | <0.01 | <0.01 | <0.01 | **2.00** |
|  | Ningi | <0.01 | <0.01 | <0.01 | <0.01 | 0.34 |
|  | Shira | <0.01 | <0.01 | <0.01 | <0.01 | <0.01 |
|  | Tafawa-Balewa | <0.01 | <0.01 | <0.01 | <0.01 | <0.01 |
|  | Toro | <0.01 | <0.01 | <0.01 | <0.01 | <0.01 |
|  | Warji | <0.01 | <0.01 | <0.01 | <0.01 | <0.01 |
|  | Zaki | **4.76** | <0.01 | <0.01 | <0.01 | <0.01 |
| Borno | Abadam | <0.01 | <0.01 | <0.01 | <0.01 | <0.01 |
|  | Askira/Uba | <0.01 | <0.01 | <0.01 | <0.01 | <0.01 |
|  | Bama | <0.01 | <0.01 | <0.01 | <0.01 | <0.01 |
|  | Bayo | <0.01 | <0.01 | <0.01 | **11.48** | <0.01 |
|  | Biu | <0.01 | <0.01 | <0.01 | <0.01 | <0.01 |
|  | Chibok | <0.01 | <0.01 | <0.01 | <0.01 | <0.01 |
|  | Damboa | <0.01 | <0.01 | <0.01 | <0.01 | <0.01 |
|  | Dikwa | <0.01 | <0.01 | <0.01 | <0.01 | <0.01 |
|  | Gubio | <0.01 | <0.01 | <0.01 | <0.01 | <0.01 |
|  | Guzamala | <0.01 | <0.01 | <0.01 | <0.01 | <0.01 |
|  | Gwoza | <0.01 | <0.01 | <0.01 | <0.01 | <0.01 |
|  | Hawul | <0.01 | <0.01 | <0.01 | <0.01 | <0.01 |
|  | Jere | <0.01 | <0.01 | <0.01 | <0.01 | **4.03** |
|  | Kaga | <0.01 | <0.01 | <0.01 | <0.01 | <0.01 |
|  | Kala/Balge | <0.01 | <0.01 | <0.01 | <0.01 | <0.01 |
|  | Konduga | <0.01 | <0.01 | <0.01 | <0.01 | <0.01 |
|  | Kukawa | <0.01 | <0.01 | <0.01 | <0.01 | <0.01 |
|  | Kwaya Kusar | <0.01 | <0.01 | <0.01 | <0.01 | <0.01 |
|  | Mafa | <0.01 | <0.01 | <0.01 | <0.01 | <0.01 |
|  | Magumeri | <0.01 | <0.01 | <0.01 | <0.01 | <0.01 |
|  | Maiduguri | <0.01 | <0.01 | <0.01 | <0.01 | <0.01 |
|  | Marte | <0.01 | <0.01 | <0.01 | <0.01 | <0.01 |
|  | Mobbar | <0.01 | <0.01 | <0.01 | <0.01 | <0.01 |
|  | Monguno | <0.01 | <0.01 | <0.01 | <0.01 | <0.01 |
|  | Ngala | <0.01 | <0.01 | <0.01 | <0.01 | <0.01 |
|  | Nganzai | <0.01 | <0.01 | <0.01 | <0.01 | <0.01 |
|  | Shani | <0.01 | <0.01 | <0.01 | <0.01 | <0.01 |
| Jigawa | Auyo | <0.01 | <0.01 | <0.01 | <0.01 | <0.01 |
|  | Babura | <0.01 | <0.01 | <0.01 | <0.01 | <0.01 |
|  | Biriniwa | <0.01 | <0.01 | <0.01 | <0.01 | <0.01 |
|  | Birnin Kudu | <0.01 | <0.01 | <0.01 | <0.01 | <0.01 |
|  | Buji | <0.01 | <0.01 | <0.01 | <0.01 | <0.01 |
|  | Dutse | <0.01 | <0.01 | <0.01 | <0.01 | <0.01 |
|  | Gagarawa | <0.01 | <0.01 | <0.01 | <0.01 | <0.01 |
|  | Garki | <0.01 | <0.01 | <0.01 | <0.01 | <0.01 |
|  | Gumel | <0.01 | <0.01 | <0.01 | <0.01 | <0.01 |
|  | Guri | <0.01 | <0.01 | <0.01 | <0.01 | **10.18** |
|  | Gwaram | <0.01 | <0.01 | <0.01 | <0.01 | <0.01 |
|  | Gwiwa | <0.01 | <0.01 | <0.01 | <0.01 | <0.01 |
|  | Hadejia | <0.01 | <0.01 | <0.01 | <0.01 | <0.01 |
|  | Jahun | <0.01 | **5.86** | <0.01 | <0.01 | <0.01 |
|  | Kafin Hausa | <0.01 | <0.01 | **9.31** | <0.01 | <0.01 |
|  | Kaugama | <0.01 | <0.01 | <0.01 | <0.01 | <0.01 |
|  | Kazaure | <0.01 | <0.01 | <0.01 | <0.01 | <0.01 |
|  | Kiri Kasama | <0.01 | <0.01 | <0.01 | <0.01 | <0.01 |
|  | Kiyawa | <0.01 | <0.01 | <0.01 | <0.01 | <0.01 |
|  | Maigatari | <0.01 | <0.01 | <0.01 | <0.01 | <0.01 |
|  | Malam Madori | <0.01 | <0.01 | <0.01 | <0.01 | <0.01 |
|  | Miga | <0.01 | <0.01 | <0.01 | <0.01 | <0.01 |
|  | Ringim | <0.01 | <0.01 | <0.01 | <0.01 | <0.01 |
|  | Roni | <0.01 | <0.01 | <0.01 | <0.01 | <0.01 |
|  | Sule Tankarkar | <0.01 | <0.01 | <0.01 | <0.01 | <0.01 |
|  | Taura | <0.01 | <0.01 | <0.01 | <0.01 | <0.01 |
|  | Yankwashi | <0.01 | <0.01 | <0.01 | <0.01 | <0.01 |
| Kaduna | Birnin-Gwari | <0.01 | <0.01 | <0.01 | <0.01 | <0.01 |
|  | Chikun | <0.01 | <0.01 | <0.01 | <0.01 | <0.01 |
|  | Giwa | <0.01 | <0.01 | <0.01 | <0.01 | <0.01 |
|  | Igabi | <0.01 | <0.01 | <0.01 | <0.01 | **2.47** |
|  | Ikara | <0.01 | <0.01 | <0.01 | <0.01 | <0.01 |
|  | Jaba | <0.01 | <0.01 | <0.01 | <0.01 | <0.01 |
|  | Jema'a | <0.01 | <0.01 | <0.01 | <0.01 | <0.01 |
|  | Kachia | <0.01 | <0.01 | <0.01 | <0.01 | <0.01 |
|  | Kaduna North | **2.03** | <0.01 | <0.01 | <0.01 | 0.59 |
|  | Kaduna South | <0.01 | **3.13** | <0.01 | <0.01 | 0.26 |
|  | Kagarko | <0.01 | <0.01 | <0.01 | <0.01 | <0.01 |
|  | Kajuru | <0.01 | <0.01 | <0.01 | <0.01 | <0.01 |
|  | Kaura | <0.01 | <0.01 | <0.01 | <0.01 | <0.01 |
|  | Kauru | <0.01 | <0.01 | <0.01 | <0.01 | <0.01 |
|  | Kubau | <0.01 | <0.01 | <0.01 | <0.01 | <0.01 |
|  | Kudan | <0.01 | <0.01 | <0.01 | <0.01 | **1.53** |
|  | Lere | <0.01 | <0.01 | <0.01 | <0.01 | <0.01 |
|  | Makarfi | <0.01 | <0.01 | <0.01 | <0.01 | <0.01 |
|  | Sabon-Gari | <0.01 | <0.01 | <0.01 | <0.01 | <0.01 |
|  | Sanga | <0.01 | <0.01 | <0.01 | <0.01 | <0.01 |
|  | Soba | <0.01 | <0.01 | <0.01 | <0.01 | <0.01 |
|  | Zangon-Kataf | <0.01 | <0.01 | <0.01 | <0.01 | <0.01 |
|  | Zaria | <0.01 | <0.01 | **5.61** | <0.01 | 0.78 |
| Kano | Ajingi | <0.01 | <0.01 | <0.01 | <0.01 | <0.01 |
|  | Albasu | <0.01 | <0.01 | <0.01 | <0.01 | <0.01 |
|  | Bagwai | <0.01 | <0.01 | <0.01 | <0.01 | <0.01 |
|  | Bebeji | <0.01 | <0.01 | <0.01 | **2.95** | <0.01 |
|  | Bichi | <0.01 | <0.01 | <0.01 | <0.01 | <0.01 |
|  | Bunkure | <0.01 | **7.25** | <0.01 | <0.01 | <0.01 |
|  | Dala | <0.01 | <0.01 | <0.01 | <0.01 | <0.01 |
|  | Dambatta | <0.01 | **5.93** | <0.01 | <0.01 | <0.01 |
|  | Dawakin Kudu | <0.01 | <0.01 | <0.01 | <0.01 | <0.01 |
|  | Dawakin Tofa | <0.01 | <0.01 | <0.01 | <0.01 | <0.01 |
|  | Doguwa | <0.01 | <0.01 | <0.01 | **3.79** | <0.01 |
|  | Fagge | <0.01 | <0.01 | <0.01 | <0.01 | <0.01 |
|  | Gabasawa | <0.01 | <0.01 | <0.01 | <0.01 | <0.01 |
|  | Garko | <0.01 | **7.79** | <0.01 | <0.01 | <0.01 |
|  | Garum Mallam | <0.01 | <0.01 | <0.01 | <0.01 | <0.01 |
|  | Gaya | <0.01 | <0.01 | <0.01 | <0.01 | <0.01 |
|  | Gezawa | <0.01 | <0.01 | <0.01 | <0.01 | <0.01 |
|  | Gwale | <0.01 | <0.01 | <0.01 | <0.01 | <0.01 |
|  | Gwarzo | <0.01 | <0.01 | <0.01 | <0.01 | 0.57 |
|  | Kabo | <0.01 | <0.01 | <0.01 | <0.01 | **3.46** |
|  | Kano Municipal | **3.97** | <0.01 | <0.01 | <0.01 | 0.57 |
|  | Karaye | <0.01 | <0.01 | <0.01 | <0.01 | <0.01 |
|  | Kibiya | <0.01 | <0.01 | <0.01 | <0.01 | <0.01 |
|  | Kiru | <0.01 | <0.01 | <0.01 | **2.15** | **3.16** |
|  | Kumbotso | <0.01 | <0.01 | <0.01 | **1.93** | **3.23** |
|  | Kunchi | <0.01 | <0.01 | <0.01 | <0.01 | <0.01 |
|  | Kura | <0.01 | <0.01 | <0.01 | <0.01 | <0.01 |
|  | Madobi | <0.01 | <0.01 | <0.01 | **4.15** | 0.78 |
|  | Makoda | <0.01 | <0.01 | <0.01 | <0.01 | <0.01 |
|  | Minjibir | <0.01 | <0.01 | <0.01 | <0.01 | **1.44** |
|  | Nasarawa | <0.01 | <0.01 | <0.01 | <0.01 | 0.71 |
|  | Rano | <0.01 | <0.01 | <0.01 | <0.01 | **3.56** |
|  | Rimin Gado | <0.01 | <0.01 | <0.01 | <0.01 | <0.01 |
|  | Rogo | <0.01 | <0.01 | <0.01 | <0.01 | <0.01 |
|  | Shanono | <0.01 | <0.01 | <0.01 | <0.01 | <0.01 |
|  | Sumaila | <0.01 | <0.01 | <0.01 | <0.01 | <0.01 |
|  | Takai | <0.01 | <0.01 | <0.01 | <0.01 | <0.01 |
|  | Tarauni | <0.01 | <0.01 | <0.01 | <0.01 | <0.01 |
|  | Tofa | <0.01 | <0.01 | <0.01 | <0.01 | <0.01 |
|  | Tsanyawa | <0.01 | <0.01 | <0.01 | <0.01 | <0.01 |
|  | Tudun Wada | <0.01 | <0.01 | <0.01 | <0.01 | <0.01 |
|  | Ungogo | **2.02** | <0.01 | <0.01 | <0.01 | **1.75** |
|  | Warawa | <0.01 | <0.01 | <0.01 | <0.01 | <0.01 |
|  | Wudil | <0.01 | <0.01 | <0.01 | <0.01 | <0.01 |
| Katsina | Bakori | <0.01 | <0.01 | <0.01 | <0.01 | <0.01 |
|  | Batagarawa | <0.01 | <0.01 | <0.01 | <0.01 | <0.01 |
|  | Batsari | <0.01 | <0.01 | <0.01 | <0.01 | <0.01 |
|  | Baure | <0.01 | <0.01 | <0.01 | <0.01 | <0.01 |
|  | Bindawa | <0.01 | <0.01 | <0.01 | <0.01 | <0.01 |
|  | Charanchi | <0.01 | <0.01 | <0.01 | <0.01 | <0.01 |
|  | Dan Musa | <0.01 | <0.01 | <0.01 | <0.01 | <0.01 |
|  | Dandume | <0.01 | <0.01 | <0.01 | <0.01 | **6.81** |
|  | Danja | <0.01 | <0.01 | <0.01 | <0.01 | <0.01 |
|  | Daura | <0.01 | <0.01 | <0.01 | <0.01 | <0.01 |
|  | Dutsi | <0.01 | <0.01 | <0.01 | <0.01 | <0.01 |
|  | Dutsin-Ma | <0.01 | <0.01 | <0.01 | <0.01 | <0.01 |
|  | Faskari | **3.40** | <0.01 | <0.01 | <0.01 | <0.01 |
|  | Funtua | <0.01 | <0.01 | <0.01 | <0.01 | <0.01 |
|  | Ingawa | <0.01 | <0.01 | <0.01 | <0.01 | <0.01 |
|  | Jibia | <0.01 | <0.01 | <0.01 | **3.04** | <0.01 |
|  | Kafur | <0.01 | <0.01 | <0.01 | <0.01 | <0.01 |
|  | Kaita | <0.01 | <0.01 | <0.01 | <0.01 | <0.01 |
|  | Kankara | <0.01 | <0.01 | <0.01 | <0.01 | <0.01 |
|  | Kankia | <0.01 | <0.01 | <0.01 | <0.01 | <0.01 |
|  | Katsina | <0.01 | **7.04** | **6.35** | <0.01 | <0.01 |
|  | Kurfi | <0.01 | <0.01 | <0.01 | <0.01 | <0.01 |
|  | Kusada | <0.01 | <0.01 | <0.01 | <0.01 | <0.01 |
|  | Mai'adua | <0.01 | <0.01 | <0.01 | <0.01 | <0.01 |
|  | Malumfashi | <0.01 | <0.01 | <0.01 | <0.01 | **2.06** |
|  | Mani | <0.01 | <0.01 | <0.01 | <0.01 | <0.01 |
|  | Mashi | <0.01 | **6.49** | <0.01 | <0.01 | 0.55 |
|  | Matazu | <0.01 | <0.01 | <0.01 | <0.01 | <0.01 |
|  | Musawa | <0.01 | <0.01 | <0.01 | <0.01 | <0.01 |
|  | Rimi | <0.01 | <0.01 | <0.01 | **6.66** | **1.23** |
|  | Sabuwa | <0.01 | <0.01 | <0.01 | <0.01 | <0.01 |
|  | Safana | <0.01 | <0.01 | <0.01 | **2.75** | <0.01 |
|  | Sandamu | **4.78** | <0.01 | <0.01 | <0.01 | <0.01 |
|  | Zango | <0.01 | <0.01 | <0.01 | <0.01 | <0.01 |
| Kebbi | Aleiro | **8.63** | <0.01 | **26.76** | <0.01 | 1.26 |
|  | Arewa-Dandi | **6.12** | <0.01 | 0.01 | **2.36** | <0.01 |
|  | Argungu | **2.87** | <0.01 | <0.01 | **2.26** | <0.01 |
|  | Augie | <0.01 | **25.39** | 0.01 | **3.86** | **1.45** |
|  | Bagudo | **7.36** | **4.10** | <0.01 | <0.01 | 0.35 |
|  | Birnin Kebbi | **4.32** | **3.67** | <0.01 | **20.27** | **1.88** |
|  | Bunza | <0.01 | <0.01 | **14.43** | **11.02** | **4.07** |
|  | Dandi | **3.90** | **13.44** | **12.36** | <0.01 | 1.13 |
|  | Fakai | <0.01 | **8.27** | <0.01 | <0.01 | 0.68 |
|  | Gwandu | <0.01 | **6.55** | <0.01 | **3.04** | 1.12 |
|  | Jega | <0.01 | **9.98** | <0.01 | <0.01 | 0.43 |
|  | Kalgo | <0.01 | <0.01 | 0.01 | **5.27** | **2.93** |
|  | Koko/Besse | <0.01 | <0.01 | <0.01 | **5.85** | **7.05** |
|  | Maiyama | **3.36** | <0.01 | <0.01 | <0.01 | <0.01 |
|  | Ngaski | <0.01 | <0.01 | <0.01 | <0.01 | <0.01 |
|  | Sakaba | <0.01 | <0.01 | <0.01 | <0.01 | <0.01 |
|  | Shanga | **4.52** | <0.01 | <0.01 | <0.01 | <0.01 |
|  | Suru | <0.01 | <0.01 | <0.01 | **3.04** | **2.82** |
|  | Wasagu/Danko | <0.01 | <0.01 | **6.73** | **1.70** | **1.57** |
|  | Yauri | **5.74** | <0.01 | <0.01 | **4.52** | **1.65** |
|  | Zuru | <0.01 | <0.01 | **10.60** | **2.74** | **3.54** |
| Niger | Agaie | <0.01 | <0.01 | <0.01 | <0.01 | <0.01 |
|  | Agwara | <0.01 | <0.01 | <0.01 | <0.01 | <0.01 |
|  | Bida | <0.01 | <0.01 | <0.01 | <0.01 | <0.01 |
|  | Borgu | <0.01 | <0.01 | <0.01 | <0.01 | <0.01 |
|  | Bosso | <0.01 | <0.01 | <0.01 | <0.01 | <0.01 |
|  | Chanchaga | <0.01 | <0.01 | <0.01 | <0.01 | <0.01 |
|  | Edati | <0.01 | <0.01 | <0.01 | <0.01 | <0.01 |
|  | Gbako | <0.01 | <0.01 | <0.01 | <0.01 | <0.01 |
|  | Gurara | <0.01 | <0.01 | <0.01 | <0.01 | <0.01 |
|  | Katcha | <0.01 | <0.01 | <0.01 | <0.01 | <0.01 |
|  | Kontagora | **4.75** | <0.01 | <0.01 | **3.63** | <0.01 |
|  | Lapai | <0.01 | <0.01 | <0.01 | <0.01 | <0.01 |
|  | Lavun | <0.01 | <0.01 | <0.01 | <0.01 | <0.01 |
|  | Magama | <0.01 | 6.65 | <0.01 | <0.01 | <0.01 |
|  | Mariga | <0.01 | <0.01 | <0.01 | <0.01 | <0.01 |
|  | Mashegu | <0.01 | <0.01 | <0.01 | <0.01 | <0.01 |
|  | Mokwa | <0.01 | <0.01 | <0.01 | <0.01 | <0.01 |
|  | Muya | <0.01 | <0.01 | <0.01 | <0.01 | <0.01 |
|  | Paikoro | <0.01 | <0.01 | <0.01 | <0.01 | <0.01 |
|  | Rafi | <0.01 | <0.01 | <0.01 | <0.01 | <0.01 |
|  | Rijau | <0.01 | **6.88** | <0.01 | <0.01 | <0.01 |
|  | Shiroro | <0.01 | <0.01 | <0.01 | <0.01 | **2.20** |
|  | Suleja | <0.01 | <0.01 | <0.01 | <0.01 | <0.01 |
|  | Tafa | <0.01 | <0.01 | <0.01 | <0.01 | <0.01 |
|  | Wushishi | <0.01 | <0.01 | <0.01 | <0.01 | <0.01 |
| Sokoto | Binji | **15.45** | **8.68** | 0.01 | **7.98** | 0.74 |
|  | Bodinga | <0.01 | <0.01 | **9.34** | **16.66** | **1.33** |
|  | Dange Shuni | **11.11** | **9.43** | **8.48** | **2.13** | **2.82** |
|  | Gada | **25.86** | **14.84** | **6.64** | **8.43** | **9.34** |
|  | Goronyo | **11.77** | **15.08** | **27.27** | **9.21** | **11.04** |
|  | Gudu | **5.67** | <0.01 | <0.01 | <0.01 | <0.01 |
|  | Gwadabawa | **13.95** | <0.01 | <0.01 | **18.07** | **7.32** |
|  | Illela | **10.72** | **18.34** | **44.22** | <0.01 | **20.13** |
|  | Isa | **7.12** | **12.09** | <0.01 | **2.75** | **7.73** |
|  | Kebbe | <0.01 | <0.01 | <0.01 | **3.38** | <0.01 |
|  | Kware | **4.01** | <0.01 | **12.39** | **12.51** | **4.62** |
|  | Rabah | **7.20** | <0.01 | <0.01 | **5.64** | **6.77** |
|  | Sabon Birni | **13.02** | **13.34** | **7.96** | **8.08** | **9.33** |
|  | Shagari | **3.44** | <0.01 | <0.01 | **5.32** | **8.37** |
|  | Silame | **5.15** | **8.69** | <0.01 | <0.01 | **2.96** |
|  | Sokoto North | **2.30** | **3.90** | **7.09** | **21.54** | **10.97** |
|  | Sokoto South | **13.57** | **4.57** | **8.39** | <0.01 | **3.53** |
|  | Tambuwal | <0.01 | <0.01 | **7.27** | **7.43** | **6.19** |
|  | Tangaza | **14.03** | **8.01** | **14.45** | **3.60** | **12.20** |
|  | Tureta | <0.01 | **39.95** | 0.01 | <0.01 | **2.25** |
|  | Wamako | **14.95** | **10.20** | **9.18** | **16.31** | **13.81** |
|  | Wurno | **6.60** | <0.01 | <0.01 | <0.01 | 0.46 |
|  | Yabo | **4.65** | <0.01 | <0.01 | <0.01 | **6.69** |
| Yobe | Bade | <0.01 | <0.01 | <0.01 | <0.01 | **17.55** |
|  | Bursari | <0.01 | <0.01 | <0.01 | <0.01 | **3.24** |
|  | Damaturu | <0.01 | <0.01 | <0.01 | **7.13** | <0.01 |
|  | Fika | <0.01 | <0.01 | <0.01 | <0.01 | <0.01 |
|  | Fune | <0.01 | <0.01 | <0.01 | <0.01 | <0.01 |
|  | Geidam | <0.01 | <0.01 | <0.01 | <0.01 | <0.01 |
|  | Gujba | <0.01 | <0.01 | <0.01 | <0.01 | <0.01 |
|  | Gulani | <0.01 | <0.01 | <0.01 | <0.01 | <0.01 |
|  | Jakusko | <0.01 | <0.01 | <0.01 | <0.01 | <0.01 |
|  | Karasuwa | <0.01 | <0.01 | <0.01 | <0.01 | <0.01 |
|  | Machina | <0.01 | <0.01 | <0.01 | <0.01 | **1.90** |
|  | Nangere | <0.01 | <0.01 | <0.01 | <0.01 | <0.01 |
|  | Nguru | <0.01 | <0.01 | **16.58** | <0.01 | <0.01 |
|  | Potiskum | <0.01 | <0.01 | <0.01 | <0.01 | <0.01 |
|  | Tarmua | <0.01 | <0.01 | <0.01 | <0.01 | <0.01 |
|  | Yunusari | <0.01 | <0.01 | <0.01 | <0.01 | <0.01 |
|  | Yusufari | <0.01 | <0.01 | <0.01 | <0.01 | <0.01 |
| Zamfara | Anka | <0.01 | <0.01 | <0.01 | <0.01 | **3.04** |
|  | Bakura | <0.01 | **5.55** | <0.01 | **5.08** | 0.94 |
|  | Birnin Magaji | **6.58** | <0.01 | <0.01 | **5.12** | 0.95 |
|  | Bukkuyum | **5.58** | <0.01 | <0.01 | <0.01 | **1.63** |
|  | Bungudu | <0.01 | **4.00** | <0.01 | **3.62** | **1.69** |
|  | Gummi | **5.92** | **5.01** | **9.05** | <0.01 | **5.51** |
|  | Gusau | **1.58** | <0.01 | <0.01 | 1.22 | **3.88** |
|  | Kaura Namoda | <0.01 | **3.66** | <0.01 | <0.01 | <0.01 |
|  | Maradun | **2.91** | <0.01 | <0.01 | <0.01 | **1.68** |
|  | Maru | <0.01 | **3.54** | <0.01 | **9.68** | **1.49** |
|  | Shinkafi | <0.01 | <0.01 | <0.01 | <0.01 | **1.92** |
|  | Talata Mafara | **2.79** | **4.74** | **17.39** | <0.01 | **4.89** |
|  | Tsafe | **2.26** | <0.01 | **6.90** | **1.77** | **3.27** |
|  | Zurmi | <0.01 | <0.01 | <0.01 | <0.01 | **1.49** |

**Supplementary Figure 1**

Trace plots indicating Markov chain convergence of the log-noma incidence risk estimates for ten LGAs.

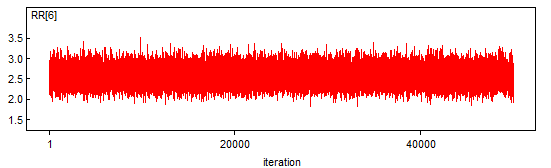

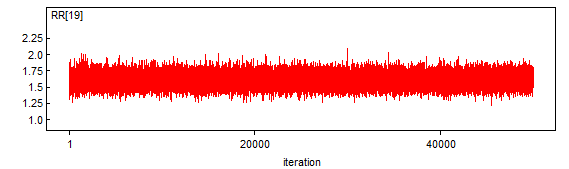

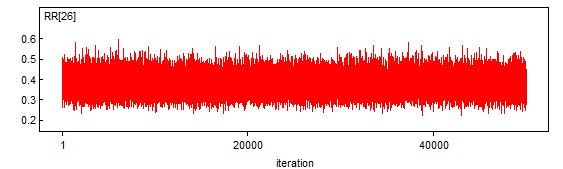

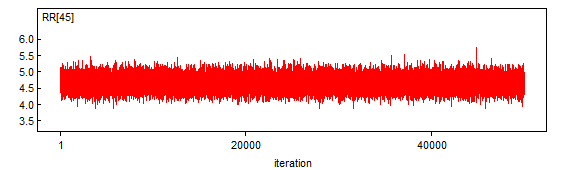

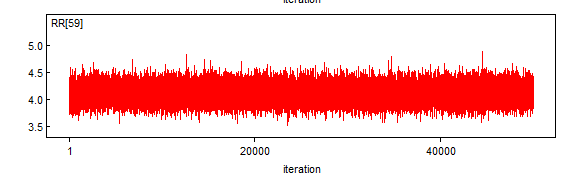

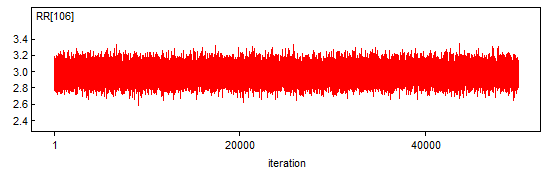

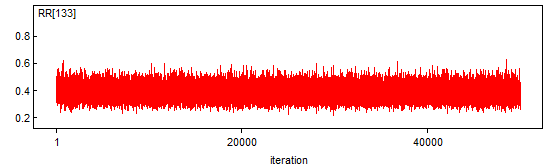

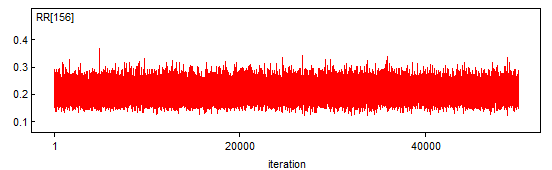

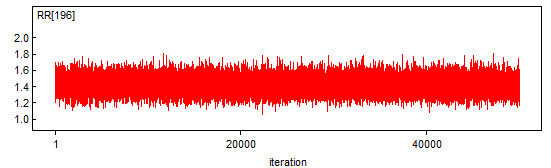

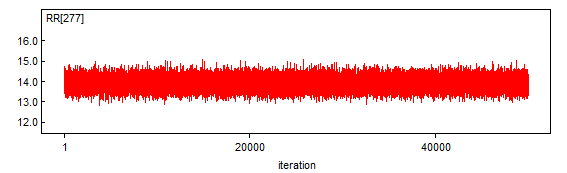

**Supplementary Figure 2**

Density plots indicating Markov chain convergence of the log-noma incidence risk estimates for ten LGAs.

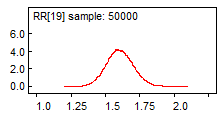

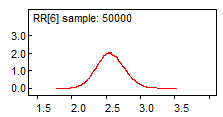

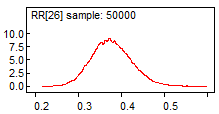

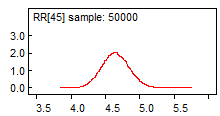

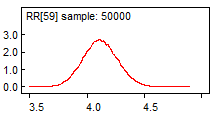

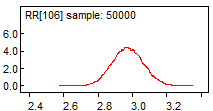

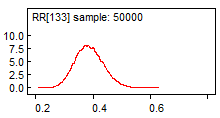

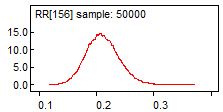

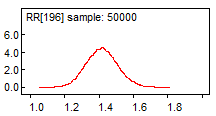

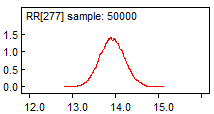

**Supplementary Figure 3**

Autocorrelation plots indicating Markov chain convergence of the log-noma incidence risk estimates for ten LGAs.

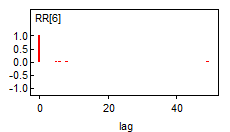

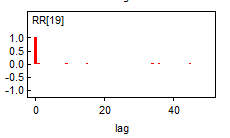

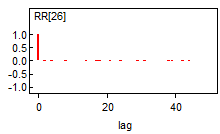

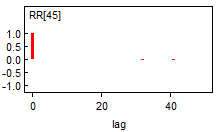

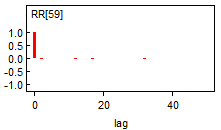

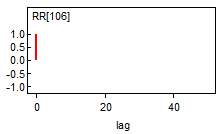

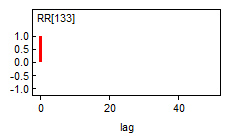

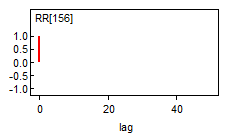

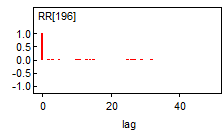

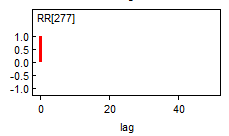

**Supplementary Figure 4**

Percentage of LGAs with a significant median SIR above average SIR in 12 Northern Nigerian states between 1999 to 2024

Percentage of LGAs with median SIR>1.01

| Northern Nigerian states | Percent of LGAs with SIR>1.01 |
| --- | --- |
| Adamawa | 4.7619 |
| Bauchi | 10 |
| Borno | 7.40741 |
| Jigawa | 3.7037 |
| Kaduna | 4.34783 |
| Kano | 11.3636 |
| Katsina | 8.82353 |
| Kebbi | 61.9048 |
| Niger | 4 |
| Sokoto | 86.9565 |
| Yobe | 17.6471 |
| Zamfara | 85.7143 |

**Supplementary Figure 5**

Line plots showing the temporal distribution of median SIR estimates at different time intervals within the study period for pertinent LGAs across 12 Northern Nigerian states. (Only LGAs with at least one median SIR estimate **above** 0.01 at any time interval were reported and line plots of LGAs below 0.01 for all time intervals were not drawn).

Adamawa

Median SIR

|  | Girei | Yola North |
| --- | --- | --- |
| 1999-2004 | 6.825e-06 | 6.349 |
| 2005-2009 | 0.0002251 | 0.0001712 |
| 2010-2014 | 0.0006367 | 0.0005485 |
| 2015-2019 | 4.66e-06 | 2.623e-06 |
| 2020-2024 | 2.809 | 8.757e-06 |

Bauchi

Median SIR

|  | Bauchi | Damban | Katagum | Kirfi | Misau | Ningi | Zaki |
| --- | --- | --- | --- | --- | --- | --- | --- |
| 1999-2004 | 4.717e-06 | 1.077e-05 | 1.031e-05 | 5.747e-06 | 4.704e-06 | 3.461e-06 | 4.756 |
| 2005-2009 | 0.0002566 | 0.0003983 | 0.0002257 | 0.0003627 | 0.0003298 | 0.000288 | 0.0003242 |
| 2010-2014 | 0.0005394 | 0.001162 | 0.0009594 | 0.001061 | 0.0008157 | 0.0006611 | 0.001241 |
| 2015-2019 | 4.733e-06 | 1.111e-05 | 2.423 | 9.336e-06 | 9.976e-06 | 3.864e-06 | 9.098e-06 |
| 2020-2024 | 1.072 | 3.515 | 0.8952 | 0.8898 | 2.001 | 0.3359 | 5.308e-06 |

Borno

Median SIR

|  | Bayo | Jere |
| --- | --- | --- |
| 1999-2004 | 7.497e-06 | 1.882e-06 |
| 2005-2009 | 0.0003525 | 0.0001902 |
| 2010-2014 | 0.0007631 | 0.001174 |
| 2015-2019 | 11.48 | 4.905e-06 |
| 2020-2024 | 9.452e-06 | 4.029 |

Jigawa

Median SIR

|  | Guri | Jahun | Kafin Hausa |
| --- | --- | --- | --- |
| 1999-2004 | 8.477e-06 | 3.926e-06 | 1.16e-05 |
| 2005-2009 | 0.0003265 | 5.864 | 0.0006352 |
| 2010-2014 | 0.001353 | 0.001284 | 9.305 |
| 2015-2019 | 8.646e-06 | 5.413e-06 | 5.363e-06 |
| 2020-2024 | 10.18 | 7.736e-06 | 7.064e-06 |

Kaduna

Median SIR

|  | Igabi | Kaduna North | Kaduna South | Kudan | Zaria |
| --- | --- | --- | --- | --- | --- |
| 1999-2004 | 1.301e-05 | 2.031 | 7.653e-06 | 1.762e-05 | 7.158e-06 |
| 2005-2009 | 0.0002946 | 0.0004006 | 3.126 | 0.0003212 | 0.0004832 |
| 2010-2014 | 0.001134 | 0.0009752 | 0.0008782 | 0.001602 | 5.614 |
| 2015-2019 | 8.413e-06 | 7.158e-06 | 1.982e-06 | 1.311e-05 | 9.953e-06 |
| 2020-2024 | 2.471 | 0.5865 | 0.2607 | 1.531 | 0.7787 |

Kano

Median SIR

|  | Bunkure | Doguwa | Garko | Gwarzo | Kabo | Kiru | Kumbotso | Madobi | Minjibir | Nasarawa | Rano | Ungogo |
| --- | --- | --- | --- | --- | --- | --- | --- | --- | --- | --- | --- | --- |
| 1999-2004 | 3.631e-06 | 8.342e-06 | 1.362e-05 | 5.969e-06 | 1.497e-05 | 1.063e-05 | 4.91e-06 | 3.204e-06 | 3.201e-06 | 7.55e-06 | 4.976e-06 | 2.016 |
| 2005-2009 | 7.253 | 0.000499 | 7.787 | 0.0004908 | 0.0002271 | 0.0002918 | 0.0002472 | 0.0002883 | 0.0002515 | 0.0001781 | 0.0005563 | 0.0001693 |
| 2010-2014 | 0.0007978 | 0.0009461 | 0.0009468 | 0.001328 | 0.001016 | 0.0008371 | 0.0008826 | 0.0009389 | 0.0008836 | 0.0003832 | 0.001047 | 0.0007141 |
| 2015-2019 | 1.536e-05 | 3.785 | 7.868e-06 | 5.732e-06 | 1.216e-05 | 2.149 | 1.925 | 4.146 | 1.739e-05 | 5.609e-06 | 1.517e-05 | 7.403e-06 |
| 2020-2024 | 2.149e-05 | 1.221e-05 | 1.007e-05 | 0.5706 | 3.458 | 3.16 | 3.227 | 0.7777 | 1.44 | 0.707 | 3.561 | 1.745 |

Katsina

Median SIR

|  | Dandume | Faskari | Jibia | Katsina | Malumfashi | Mashi | Rimi | Safana | Sandamu |
| --- | --- | --- | --- | --- | --- | --- | --- | --- | --- |
| 1999-2004 | 1.519e-05 | 3.398 | 1.049e-05 | 3.195e-06 | 3.311e-06 | 1.163e-05 | 1.233e-05 | 1.021e-05 | 4.779 |
| 2005-2009 | 0.0007745 | 0.0006314 | 0.0003366 | 7.037 | 0.0003635 | 6.492 | 0.000581 | 0.0009807 | 0.0009458 |
| 2010-2014 | 0.001684 | 0.001987 | 0.001026 | 6.348 | 0.001273 | 0.001486 | 0.001274 | 0.00145 | 0.0008714 |
| 2015-2019 | 9.812e-06 | 1.313e-05 | 3.037 | 8.483e-06 | 7.58e-06 | 1.006e-05 | 6.662 | 2.754 | 5.517e-06 |
| 2020-2024 | 6.809 | 1.137e-05 | 6.435e-06 | 4.124e-06 | 2.057 | 0.5505 | 1.225 | 2.184e-05 | 1.145e-05 |

Kebbi

Median SIR

|  | Aleiro | Arewa-Dandi | Argungu | Augie | Bagudo | Birnin Kebbi | Bunza | Dandi | Fakai | Gwandu | Jega | Kalgo | Koko/Besse | Maiyama | Shanga | Suru | Wasagu/Danko | Yauri | Zuru |
| --- | --- | --- | --- | --- | --- | --- | --- | --- | --- | --- | --- | --- | --- | --- | --- | --- | --- | --- | --- |
| 1999-2004 | 8.632 | 6.118 | 2.865 | 2.283e-05 | 7.362 | 4.318 | 3.124e-05 | 3.899 | 1.724e-05 | 2.022e-05 | 1.652e-05 | 4.491e-05 | 2.153e-05 | 3.36 | 4.516 | 1.897e-05 | 8.902e-06 | 5.742 | 1.109e-05 |
| 2005-2009 | 0.001449 | 0.0006948 | 0.0004823 | 25.39 | 4.099 | 3.665 | 0.0007993 | 13.44 | 8.266 | 6.553 | 9.981 | 0.001128 | 0.000634 | 0.0005099 | 0.001945 | 0.0006058 | 0.0004845 | 0.00067 | 0.0003902 |
| 2010-2014 | 26.76 | 0.005767 | 0.002698 | 0.006123 | 0.001795 | 0.0009029 | 14.43 | 12.36 | 0.004726 | 0.003589 | 0.00279 | 0.005669 | 0.003835 | 0.002965 | 0.003195 | 0.002818 | 6.727 | 0.001525 | 10.6 |
| 2015-2019 | 5.118e-05 | 2.363 | 2.262 | 3.862 | 1.866e-05 | 20.27 | 11.02 | 1.569e-05 | 1.172e-05 | 3.038 | 2.163e-05 | 5.271 | 5.849 | 2.488e-05 | 2.138e-05 | 3.036 | 1.699 | 4.515 | 2.741 |
| 2020-2024 | 1.262 | 4.173e-05 | 3.339e-05 | 1.452 | 0.3486 | 1.875 | 4.072 | 1.133 | 0.6834 | 1.118 | 0.425 | 2.93 | 7.048 | 1.927e-05 | 2.159e-05 | 2.817 | 1.573 | 1.648 | 3.539 |

Niger

|  | Kontagora | Magama | Rijau | Shiroro |
| --- | --- | --- | --- | --- |
| 1999-2004 | 4.754 | 6.27e-06 | 1.126e-05 | 1.407e-05 |
| 2005-2009 | 0.0005699 | 6.652 | 6.875 | 0.0002931 |
| 2010-2014 | 0.002256 | 0.001866 | 0.002976 | 0.001066 |
| 2015-2019 | 3.629 | 1.276e-05 | 1.109e-05 | 6.047e-06 |
| 2020-2024 | 9.326e-06 | 1.988e-05 | 2.02e-05 | 2.196 |

Sokoto

Median SIR

|  | Binji | Bodinga | Dange Shuni | Gada | Goronyo | Gudu | Gwadabawa | Illela | Isa | Kebbe | Kware | Rabah | Sabon Birni | Shagari | Silame | Sokoto North | Sokoto South | Tambuwal | Tangaza | Tureta |
| --- | --- | --- | --- | --- | --- | --- | --- | --- | --- | --- | --- | --- | --- | --- | --- | --- | --- | --- | --- | --- |
| 1999-2004 | 15.45 | 2.091e-05 | 11.11 | 25.86 | 11.77 | 5.668 | 13.95 | 10.72 | 7.115 | 1.308e-05 | 4.012 | 7.195 | 13.02 | 3.438 | 5.147 | 2.3 | 13.57 | 1.741e-05 | 14.03 | 5.92e-05 |
| 2005-2009 | 8.679 | 0.0006615 | 9.428 | 14.84 | 15.08 | 0.002094 | 0.0006306 | 18.34 | 12.09 | 0.0005907 | 0.001203 | 0.0009184 | 13.34 | 0.0008324 | 8.693 | 3.902 | 4.566 | 0.000942 | 8.005 | 39.95 |
| 2010-2014 | 0.006214 | 9.338 | 8.475 | 6.635 | 27.27 | 0.004969 | 0.003868 | 44.22 | 0.003926 | 0.003825 | 12.39 | 0.004812 | 7.963 | 0.004698 | 0.00459 | 7.088 | 8.394 | 7.266 | 14.45 | 0.007329 |
| 2015-2019 | 7.976 | 16.66 | 2.132 | 8.429 | 9.206 | 4.613e-05 | 18.07 | 2.392e-05 | 2.754 | 3.384 | 12.51 | 5.636 | 8.083 | 5.321 | 1.209e-05 | 21.54 | 1.195e-05 | 7.429 | 3.6 | 2.97e-05 |
| 2020-2024 | 0.7393 | 1.325 | 2.819 | 9.338 | 11.04 | 3.159e-05 | 7.32 | 20.13 | 7.733 | 2.633e-05 | 4.619 | 6.774 | 9.326 | 8.371 | 2.957 | 10.97 | 3.531 | 6.188 | 12.2 | 2.252 |

Yobe

Median SIR

|  | Bade | Bursari | Damaturu | Machina | Nguru |
| --- | --- | --- | --- | --- | --- |
| 1999-2004 | 1.684e-05 | 1.874e-05 | 4.267e-06 | 1.397e-05 | 1.311e-05 |
| 2005-2009 | 0.0003484 | 0.0004579 | 0.0005588 | 0.000613 | 0.000552 |
| 2010-2014 | 0.001434 | 0.001613 | 0.0007934 | 0.002468 | 16.58 |
| 2015-2019 | 1.183e-05 | 1.633e-05 | 7.133 | 1.225e-05 | 6.783e-06 |
| 2020-2024 | 17.55 | 3.238 | 7.804e-06 | 1.895 | 4.847e-06 |

Zamfara

Median SIR

|  | Anka | Bakura | Birnin Magaji | Bukkuyum | Bungudu | Gummi | Gusau | Kaura Namoda | Maradun | Maru | Shinkafi | Talata Mafara | Tsafe | Zurmi |
| --- | --- | --- | --- | --- | --- | --- | --- | --- | --- | --- | --- | --- | --- | --- |
| 1999-2004 | 3.737e-05 | 1.973e-05 | 6.579 | 5.58 | 5.357e-06 | 5.919 | 1.578 | 1.906e-05 | 2.907 | 1.358e-05 | 1.271e-05 | 2.791 | 2.264 | 1.428e-05 |
| 2005-2009 | 0.001085 | 5.545 | 0.001197 | 0.0008904 | 4.001 | 5.008 | 0.0002759 | 3.662 | 0.0003283 | 3.539 | 0.0005657 | 4.736 | 0.0004098 | 0.0003788 |
| 2010-2014 | 0.003664 | 0.00442 | 0.003844 | 0.003843 | 0.001679 | 9.05 | 0.001546 | 0.001993 | 0.002337 | 0.002261 | 0.002349 | 17.39 | 6.898 | 0.00159 |
| 2015-2019 | 2.635e-05 | 5.084 | 5.122 | 1.112e-05 | 3.622 | 1.736e-05 | 1.215 | 1.609e-05 | 1.053e-05 | 9.682 | 2.947e-05 | 1.008e-05 | 1.766 | 1.07e-05 |
| 2020-2024 | 3.042 | 0.9401 | 0.9455 | 1.631 | 1.687 | 5.508 | 3.883 | 1.035e-05 | 1.678 | 1.494 | 1.92 | 4.887 | 3.272 | 1.493 |
